# Use of routine clinical data in adults with chronic diseases for identifying risk of frailty: results from the UK Biobank study

**DOI:** 10.64898/2026.10.05.26364730

**Authors:** Benjamin Landré, Gabriella C Silva, Aurore Fayosse, Ian Meneghel Danilevicz, Wolfgang Lieb, Séverine Sabia, Archana Singh-Manoux, Paula Stürmer

## Abstract

Frailty assessment is increasingly recommended for risk stratification in chronic disease management, but systematic clinical screening for frailty remains difficult to implement because it is resource-intensive. We investigated whether routinely collected clinical measures could be repurposed to prioritize adults with chronic diseases for clinical frailty assessment.

We conducted a prediction study using UK Biobank data. A stacked ensemble model was developed using 12 routinely and low cost measured clinical variables to predict frailty. Gain metrics were used to quantify the proportion of frailty cases identified when screening was restricted to individuals with the highest predicted risk of frailty. We compared health profiles (mortality, polypharmacy, self-rated health, falls) of frail and non-frail individuals across different levels of predicted risk. The robustness of this screening strategy was evaluated across 3,284 modelling settings.

Among 281,729 participants (mean age 57·9; 54·7% women) with at least one chronic condition, 5·8% were frail. In the validation sample (N validation=93,910, N training=187,819), screening only the 20% of participants with the highest predicted risk of frailty identified 55·6% of all frailty cases, corresponding to a 2·78-fold improvement compared to random screening. Overall, the model recovered 53·4% of the maximum achievable improvement over random screening. Frailty remained always associated with poor outcomes. Screening performance remained consistent across alternative modelling settings.

Routinely collected clinical measures can improve the efficiency of frailty screening among adults living with chronic diseases by prioritizing individuals based on their level of risk. Since these data are already gathered during routine care, this risk-prioritization strategy could support scalable screening without increasing testing burden or clinical workload.

## Introduction

Frailty is a clinical state of reduced physiological reserve that increases vulnerability to adverse health outcomes following stressors^1^. The prevalence of frailty increases with age^1^ particularly among people with chronic diseases^2,3^. Across a wide range of chronic diseases, frailty is consistently associated with higher risk of poor health outcomes,^4–7^ independently of chronological age. The assessment of frailty is increasingly recommended in clinical guidelines^8–10^ for management of chronic diseases to support risk stratification and to personalize care. Because frailty is potentially reversible, timely identification is important. Multicomponent interventions such as resistance training and nutritional support improve health outcomes among frail individuals^11,12^ and is likely to be cost-effective^13^.

Translating recommendations on frailty assessment into routine practice remains challenging. Frailty can be assessed using simple tools that requires little equipment,^14^ but healthcare professionals may not always have the time to undertake the evaluation. Current screening strategies for frailty remain confined to older ages^15–18^, or disease-based despite major chronic diseases being common in midlife^19^ and frailty onset seen before old age^2^. Among people living with chronic diseases, frailty may affect a small proportion of the population^7^, making universal screening within this subgroup inefficient. Widespread screening may be more appropriate in the oldest-old, where the prevalence of frailty is higher^1^.

Routine clinical testing may offer a more efficient opportunity opportunity to identify those individuals, who should be assessed for frailty. Most individuals with chronic diseases undergo repeat blood testing over the course of their clinical care that generates large volumes of routinely collected data^20^. These data are not intended to assess frailty, and no individual clinical measure is likely to replace a,specific tool to measure frailty. Nevertheless, routine clinical data could be useful for screening for further assessment for frailty as in other contexts; for example, to choose individuals to include in clinical trials^21^. This approach would be more efficient than populational screening strategies without adding tests or increasing clinical workload^22^.

According, we examined whether 12 routinely collected clinical measures could improve the targeting of frailty screening among adults with chronic diseases in the UK Biobank study.

## Methods

### Study design and population

The UK Biobank is a prospective, population-based cohort study that includes over 500,000 participants recruited between 2006 and 2010. We used data from the baseline assessment that comprised face-to-face interviews, self-completed questionnaires, physical examinations, and the collection of biosamples. All participants provided written, informed consent for data collection, analysis, and access to their health records. UK Biobank received approval from the National Information Governance Board for Health and Social Care and the NHS North West Centre for Research Ethics Committee (reference number 11/NW/0382). This research was conducted using the UK Biobank Resource under data sharing application number 96856.

We included participants with at least one chronic disease, identified using 38 chronic diseases based on the list defined by Ronaldson et al^23^ (**Supplementary Table 1**) and used in other UK Biobank studies^2^. Chronic disease ascertainment was determined using linked electronic health records (including primary care, hospital admission, and cancer registry data). We excluded participants with missing data on frailty status, age, sex, or more than two routine clinical measures. The final analytical sample included 281,729 participants (**Supplementary Figure 1**).

### Frailty

Fried’s frailty phenotype (FFP)^24^ was defined using a measure adapted to UK Biobank data proposed by Jiang and colleagues^25^ and includes the following five components:

- Weight loss was determined based on participants’ self-reported weight change in the previous year and was categorized as reporting weight loss and no weight loss (without change or weight gain).
- Exhaustion was based on reported tiredness over the past 2 weeks, with responses including “not at all”, “over several days”, “more than half the days”, or “nearly every day”. Participants who reported tiredness on more than half the days or nearly every day were classified as meeting the criterion for exhaustion.
- Slow walking pace was based on the question “How would you describe your usual walking pace?” with responses being slow, average, or brisk. Participants who reported slow walking pace were considered to fulfil the criterion for slow walking pace.
- Physical activity was categorized into four groups based on activities in the last four weeks: none (no physical activity), low (light activities such as pruning or watering the lawn), medium (walking for pleasure or heavy activities such as weeding, carpentry, or digging), and high (strenuous sports). Participants who reported no or low activity were classified as meeting the criterion for low physical activity.
- Grip strength was measured using a hydraulic hand dynamometer on both arms. The mean of these values or the measure available was considered. The cut-offs to define low grip strength were based on sex and body mass index as defined in the FFP^24^.

The five criteria were summed to obtain a score from 0 to 5, and participants were classified as frail (score ≥ 3), or non-frail (score ≤ 2) as in the FFP.

### Routine clinical measures (measured at baseline)

We considered twelve clinical measures commonly assessed during routine management of chronic diseases. Selection of these measures was based on their widespread availability in routine clinical practice and their plausible association with frailty. These measures were:

- Anthropometric measure: Body Mass Index (BMI)
- Blood lipids: total cholesterol, low-density lipoprotein (LDL) cholesterol, high-density lipoprotein (HDL) cholesterol, triglycerides
- Markers of hepatic function: alanine aminotransferase (ALAT), aspartate aminotransferase (ASAT), gamma-glutamyl transferase (GGT)
- Markers of renal function: creatinine, estimated glomerular filtration rate (eGFR)
- Marker of glucose metabolism: glycated hemoglobin A1c (HbA1c), glucose

To provide some context, we have listed the costs of the various measures. Costs were estimated using publicly available reimbursement data from the French National Health Insurance system^26^. Measurement procedures and cost-calculation are detailed in **Supplementary Method 1**.

### Clinical profiles

To characterize the clinical profile of individuals prioritized for screening, we evaluated all-cause mortality at 3- and 10-year horizons, assessed using linkage to national death registries and cross-sectional self-reported health (falls, polypharmacy (≥5 self-reported medications), and poor overall health).

Participants with missing data on falls, polypharmacy, or overall health were assigned to the reference category. Linkage with national death registries is considered exhaustive.

### Statistical analysis

First, we examined whether routinely collected clinical measures could improve the efficiency of frailty screening among people living with chronic diseases. The frailty prediction model was developed using a stacked ensemble combining extreme gradient boosting and penalized logistic regression^27^. Age, sex, and the 12 routine clinical measures were considered as predictors. The dataset was randomly divided into stratified training (2/3) and validation (1/3) sets while maintaining the frailty prevalence observed in the full sample. All pre-processing procedures, including missing-value imputation, feature pre-processing, hyperparameters optimization, were performed within repeated stratified cross-validation to prevent information leakage (i.e., inadvertent use of validation-fold information within cross-validation) (**Supplementary Methods 2**).

Because the intended application was prioritization for frailty screening rather than diagnosis of frailty, model performance was primarily evaluated using gain and lift metrics. Gain was defined as the cumulative proportion of frail individuals identified when screening was restricted to participants with the highest predicted risk, whereas lift represented the improvement over random selection.

Gain and lift were calculated for the top k=10%, 20%, 30%, 50%, 75%, and 100% (i.e total population) of participants ranked according to predicted probability of frailty?. These thresholds were chosen to reflect a range of potential screening options, from highly targeted programs to population-wide screening. Overall performance was summarized using the normalized area under the gain curve compared with random (random selection without prioritization) and optimal (all frail participants selected first) reference models^28^. Receiver operating characteristic area under the curve (ROC-AUC) and precision-recall area under the curve (PR-AUC) were reported as secondary performance measures. Definitions and interpretation of gain metrics are summarized in **Supplementary Methods 3**.

Second, we assessed the clinical relevance of prediction prioritization in a screening setting. For each screening threshold, defined by the proportion of participants with the highest predicted risk (top k%), we examined whether observed frailty remained associated with adverse health outcomes among those prioritized for screening. We considered 3-year and 10-year mortality, falls during the previous year, polypharmacy, and self-reported poor health. Associations between observed frailty and mortality were estimated using Cox proportional hazards models, and associations with binary outcomes using logistic regression models. Results are reported as hazard ratios (HRs) and odds ratios (ORs), respectively, with 95% confidence intervals (CIs).

Third, we evaluated the stability of screening performance across alternative combinations of available routine clinical measure panels and participants’ characteristics. To accomplish this, we maintained a similar prediction procedure as that described previously with minor modifications to the parameters to achieve reasonable processing times (**Supplementary Methods 4**). The same pre-processing procedures were applied throughout. For this analysis, routine clinical tests comprising three to 12 available measures were evaluated. For each panel size, 20% of all possible combinations of routine clinical measures were randomly sampled, with a minimum of 20 combinations retained; when fewer than 20 combinations existed, all combinations were analyzed (**Supplementary Methods 5**). This resulted in 821 combinations. Stability was further assessed across age strata (<60 vs ≥60 years) and across alternative chronic disease definitions, ranging from an exhaustive list including 38 diseases to a restricted list comprising the 12 most prevalent diseases, based on the approach proposed by Fortin et al.^29^. Overall, 3,284 model variations were considered (821 combination patterns x 4 strata) (**Supplementary Methods 5, Supplementary Table 2**). To report dispersion across strata, we calculated the median, 1^st^ and 3^rd^ quartiles, interquartile range (IQR), 5% and 95% percentiles of gain and lift at top 10/20/30% as well as normalized gain.

To quantify the relative contribution of parameters to variability in model performance, we fitted a linear mixed-effects model with a random intercept for each of the 821 routine clinical measure combination patterns. The outcome was gain at top 20%, with age strata, chronic disease definition, number of clinical measures included in the combination, and combination cost entered as fixed-effect. Variance decomposition was performed using marginal and conditional R² to quantify the proportion of variance explained by fixed effects alone and by the full model, respectively. Within the fixed-effects component, predictor-specific contributions were assessed using semi-partial R² (part R²), representing the unique variance explained by each predictor, and inclusive R², accounting for both unique and shared variance among correlated predictors^30^.

Finally, to quantify the contribution of individual routine clinical measures to model performance, we compared the median gain at top 20% across panels including versus excluding each routine clinical measure, stratified by the number of routine clinical measures included in the panel. For each routine clinical measure and panel size, the difference in median gain between routine clinical measures that contained and excluded these panels was calculated.

In sensitivity analyses, given the strong relationship between frailty and age and frailty and cardio-metabolic diseases, we examined the robustness of the primary findings across four age groups defined by age quartiles and according to the presence/absence of metabolic (ICD-10 Chapter IV) and presence/absence of cardiometabolic (ICD-10 Chapter IV and/or IX) diseases.

## Results

From 501,941 participants at baseline in the UK Biobank study, 309,770 had at least one chronic disease, and 281,729 (93.2%) had data on frailty status, routine clinical measures and covariates. The analytical sample included 154,085 (54.7%) women, the mean age was 57.9 (± 7.8) years, and 5.8% (16259) participants were frail. The distribution of all measures in the training (N_2/3_= 187,819) and validation (N_1/3_ = 93,910) datasets were similar (**Supplementary Table 3**).

The validation set comprised 93,910 participants, including 5,461 (5.8%) participants classified as frail. Gain curve analyses (**Figure 1**; **Supplementary Table 4**) showed that restricting frailty screening to 10% of participants with the highest predicted risk using age, sex, and 12 routine clinical measures identified 38.1% of all frailty cases, while screening the top 20% and 30% identified 55.6% and 67.1% of cases, respectively. Correspondingly, frailty prevalence decreased from 22.1% in the top 10% (lift = 3.81) to 16.2% in the top 20% (lift = 2.78) and 13.0% in the top 30% (lift = 2.24), remaining substantially higher than the overall prevalence of 5.8%. Across these screening thresholds of the prediction model, every increase in the screened population yielded proportionally greater gains in frailty detection than would be expected from random selection. Beyond the top 30%, however, progressively larger increases in the screened population resulted proportionally in smaller increases in gain. Overall, the model achieved a normalized gain AUC of 53.4%, indicating over half of the maximum achievable improvement over random screening. Discriminative performance was moderate, with a ROC-AUC of 0.767 and a PR-AUC of 0.227.

**Figure 1.**
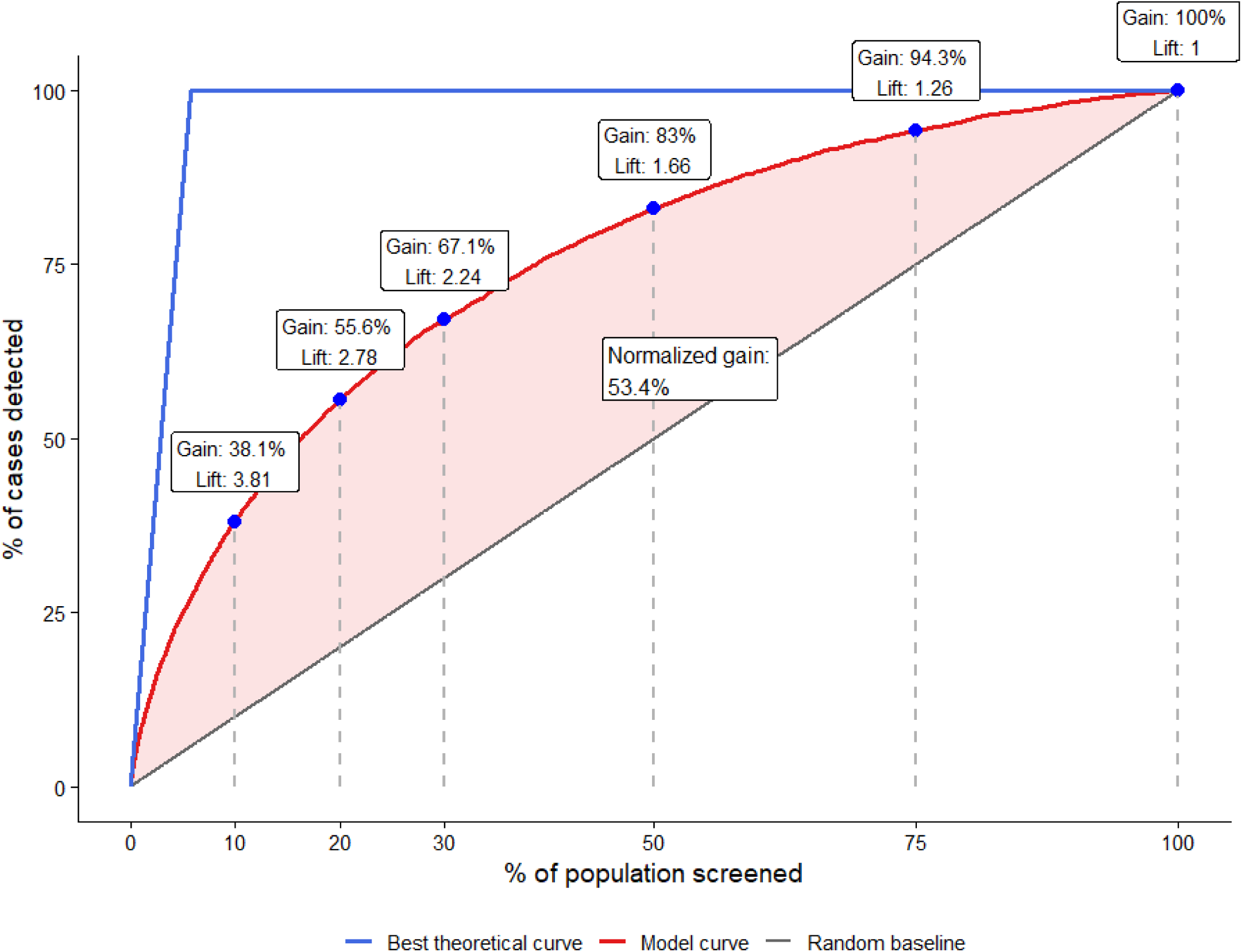
Gain curve of stack model estimated in validation sample (N = 93,910).

Within each screening stratum, observed frailty identified a subgroup with a consistently less favourable clinical profile (**Table 1**). For example, in those in the top 20% of predicted frailty, frailty was significantly associated with 3-year mortality (HR 1.90, 95% CI 1.51–2.38), 10-year mortality (HR 1.91, 1.72–2.11), poor self-rated health (OR 7.17, 6.55–7.84), polypharmacy (OR 2.99, 2.75–3.25), and falls (OR 2.40, 2.21–2.60). As progressively larger proportions of the population were included for screening, the associations between frailty and adverse outcomes became stronger. This increase was primarily driven by a greater reduction in events among individuals classified as non-frail than among those classified as frail. For example, for poor self-rated health, prevalence among frail individuals remained relatively stable (43.8% in the top 10% vs 39.6% in the complete sample), whereas prevalence among non-frail individuals decreased from 11.6% to 4.1%, resulting in an increase in the odds ratio from 5.97 to 15.45. Similar patterns were observed for all indicators, suggesting progressively greater separation between frail and non-frail individuals as the screening threshold increased.

**Table 1.** Clinical characteristics of frail participants compared to non-frail in validation sample when k% participants are screened based on their probability of being frail (N= 93,910).

|  | k% of participants screened = k% of participants with the largest predicted probability of frailty<sup>A</sup> |  |  |  |  |  |
| --- | --- | --- | --- | --- | --- | --- |
|  | k=10% | k=20% | k=30% | k=50% | k=75% | k=100% (Complete sample) |
| **% Frail** | 22.1% | 16.2% | 13.0% | 9.7% | 7.3% | 5.8% |
| **Gain** | 38.1% | 55.6% | 67.1% | 83.0% | 94.3% | 100% |
| **3-year mortality** |  |  |  |  |  |  |
| % event in frail | 3.8% | 3.4% | 3.2% | 3.0% | 2.9% | 2.8% |
| % event in non-frail | 2.0% | 1.8% | 1.6% | 1.4% | 1.2% | 1.0% |
| HR (95% CI) | 1.93 (1.47–2.53) | 1.90 (1.51–2.38) | 2.00 (1.63–2.46) | 2.17 (1.80–2.62) | 2.48 (2.08–2.96) | 2.85 (2.40–3.39) |
| **10-year mortality** |  |  |  |  |  |  |
| % event in frail | 17.7% | 16.6% | 16.2% | 15.1% | 14.6% | 14.3% |
| % event in non-frail | 10.2% | 9.1% | 8.2% | 7.2% | 6.4% | 5.7% |
| HR (95% CI) | 1.82 (1.61–2.06) | 1.91 (1.72–2.11) | 2.06 (1.88–2.26) | 2.20 (2.02–2.39) | 2.39 (2.21–2.59) | 2.64 (2.45–2.85) |
| **Poor self-rated health** |  |  |  |  |  |  |
| % event in frail | 43.8% | 42.0% | 41.1% | 40.4% | 39.6% | 39.6% |
| % event in non-frail | 11.6% | 9.2% | 7.8% | 6.1% | 4.9% | 4.1% |
| OR (95% CI) | 5.97 (5.33–6.68) | 7.17 (6.55–7.84) | 8.26 (7.65–8.96) | 10.38 (9.66–11.15) | 12.63 (11.82–13.49) | 15.45 (14.49–16.46) |
| **Polypharmacy** |  |  |  |  |  |  |
| % event in frail | 73.9% | 71.0% | 68.7% | 65.1% | 63.1% | 61.7% |
| % event in non-frail | 52.0% | 45.0% | 40.3% | 34.0% | 28.2% | 23.9% |
| OR (95% CI) | 2.61 (2.35–2.91) | 2.99 (2.75–3.25) | 3.26 (3.03–3.51) | 3.63 (3.40–3.87) | 4.35 (4.10–4.62) | 5.14 (4.86–5.44) |
| **Falls** |  |  |  |  |  |  |
| % event in frail | 46.8% | 46.1% | 46.1% | 45.0% | 44.8% | 44.7% |
| % event in non-frail | 28.0% | 26.3% | 24.9% | 23.4% | 21.6% | 20.4% |
| OR (95% CI) | 2.27 (2.05–2.51) | 2.40 (2.21–2.60) | 2.58 (2.40–2.77) | 2.69 (2.53–2.86) | 2.95 (2.78–3.12) | 3.14 (2.97–3.32) |

HRs were estimated using Cox proportional hazards models. ORs were estimated using logistic regression.
<sup>A</sup>Top k% refers to the percentage of participants, sorted by decreasing predicted frailty, included in the stratum.
Abbreviations: HR: Hazard Ratio; OR: Odds Ratio; CI: Confidence Interval.

We assessed the stability of the risk-prioritization strategy by evaluating dispersion of gain, lift, and normalized gain (**Figure 2, Supplementary Table 5**) in 3,284 alternative settings (821 combinations of clinical test panels, across 4 different population subgroups). Overall, the median normalized gain was 39.4% (IQR 34.3–42.9%, 5% percentile = 28.1%). The top 10%, 20%, and 30% highest-ranked individuals captured a median of 29.6%, 46.0%, and 57.9% of frailty cases, corresponding to median lift values of 3.0, 2.3, and 1.9, respectively. Variability across analyses was limited; at a screening threshold of 20% of the population, gain IQR varied by 7.9 percentage points (41.0–48.9%, 5th percentile = 37.0%), while nearly 95% of lift remained consistently above two-fold enrichment (IQR 2.1–2.5; at 5^th^ percentile = 1.9). These findings suggest that the risk-prioritization strategy always outperforms random screening across alternative settings.

**Figure 2.**
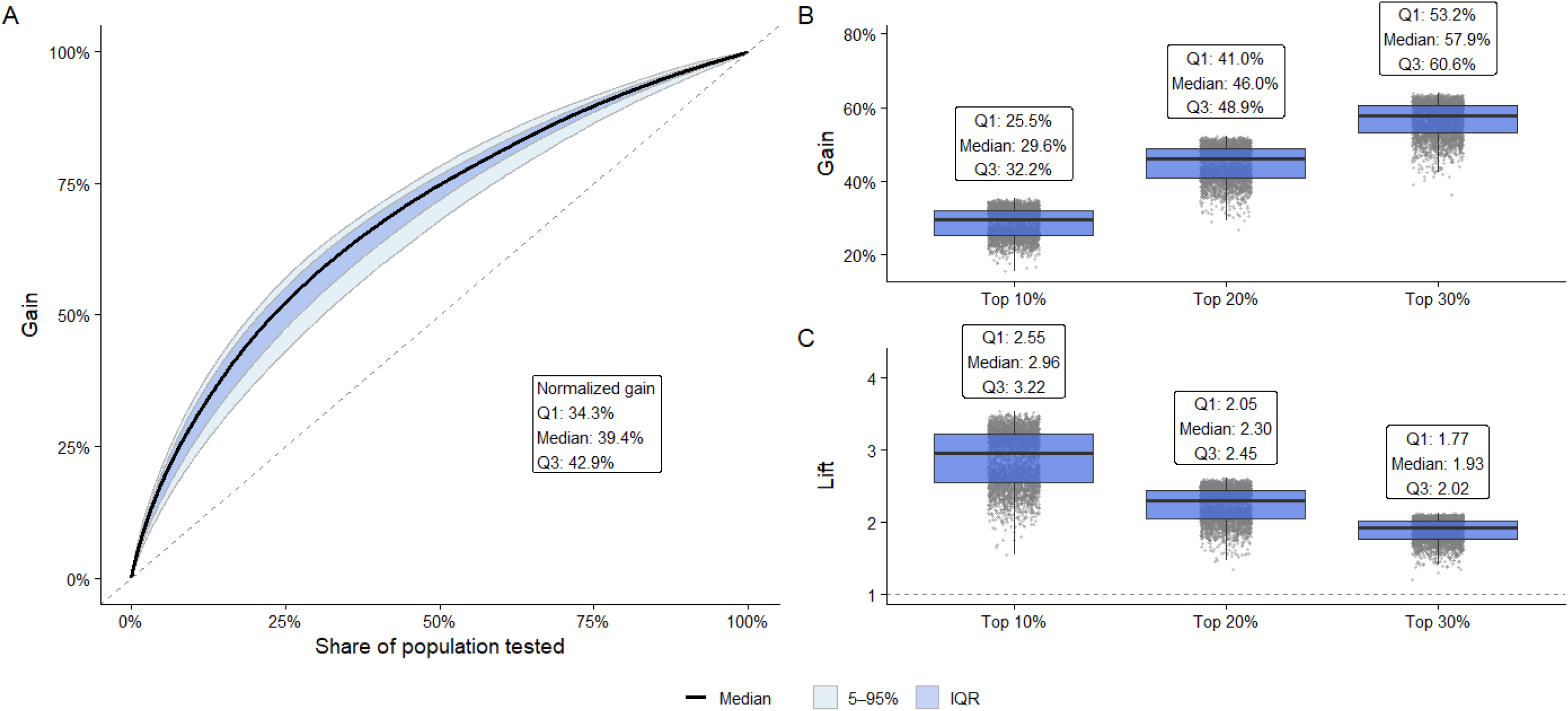
Gain curves distribution (A), gain distribution at top 10/20/30% (B), corresponding lift distribution (C) across 3,284 settings. Abbreviations: IQR: interquartile range; Q1: 1^st^ quartile; Q3: 3^rd^ quartile.

We assessed the factors contributing to gain at top 20% using mixed models (**Table 2**). The mixed-effects model explained 20.4% of the outcome variance through fixed effects, while the marginal R² was 93.0%, suggesting that the between-combination variability accounted for a major proportion of the explained variance. Among fixed effects, the number of clinical measures was the largest unique contributor, explaining 7.2% of total outcome variance (partial R²=7.2; 95% CI 2.2–12.4). Panel costs showed a substantial overall association with gain (inclusive R²=13.1; 95% CI 9.7–17.1), but contributed little unique variance after accounting for other predictors (partial R²=0.2; 95% CI 0.0– 5.6), indicating substantial overlap with other covariates, particularly the size of combination (correlation = 0.746?). Age group and choice of list contributed minimally to explained variance. For both combination size and total costs, we observed increasing gain at top 20% for increasing values of these parameters (**Supplementary Figure 2**). Conclusions remained similar for gain at top 10 or 30% (**Supplementary Table 6**).

**Table 2.** Determinants of variability in gain when participants with the top 20% of predicted risk are screened.

| Predictor | Unique variance explained (Part R<sup>2</sup>) | Total variance associated with predictor (Inclusive R<sup>2</sup>) |
| --- | --- | --- |
| Number of clinical measures | 7.2% (2.2; 12.4) | 20.1% (15.7; 24.4) |
| Panel cost | 0.2% (0.0; 5.6) | 13.1% (9.7; 17.1) |
| Age group | 0.0% (0.0; 5.6) | 0.0% (0.0; 0.0) |
| Disease list choice | 0.1% (0.0; 5.5) | 0.1% (0.1; 0.2) |
| Marginal R<sup>2</sup> = 93.0%; |  |  |
| Conditional R<sup>2</sup> = 24.0%; |  |  |
| Random intercept variance attributable to clinical-measure combination patterns = 72.6%. |  |  |

To assess the contribution of individual routine clinical measures, we compared the median top-20% gain between panels including versus excluding each of the routine clinical measures within each combination size. Routine clinical measure effects varied substantially according to panel composition (**Figure 3, Supplementary Table 7**). BMI showed the most consistent positive contribution, with median gain of approximately 6 percentage points across several combination sizes. In contrast, most routine clinical measures had context-dependent effects, improving performance in some panel configurations but not others. Overall, routine clinical measures-specific effects tended to attenuate as combination size increased, suggesting increasing redundancy among routine clinical measures in larger panels.

**Figure 3.**
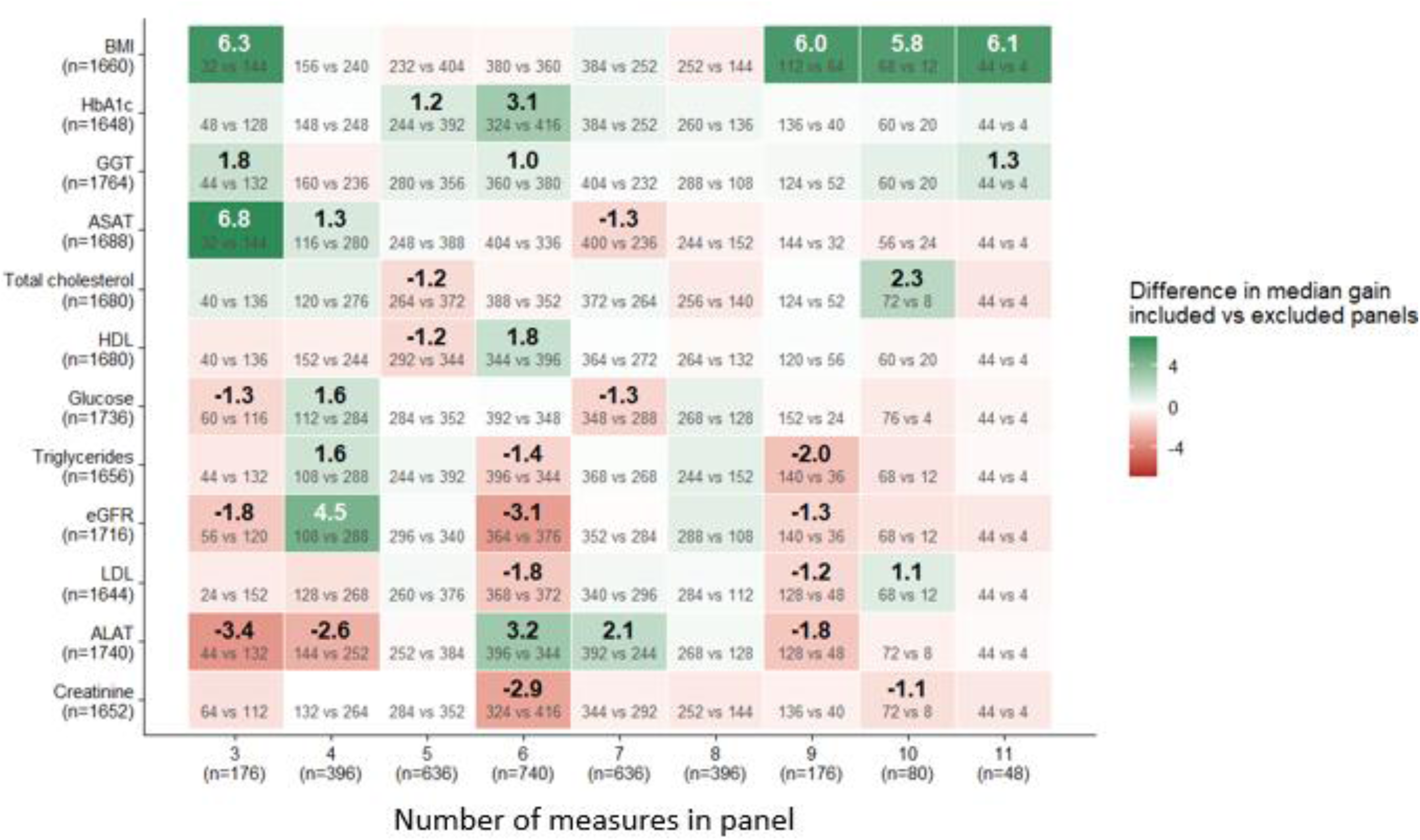
Difference* in median gain when participants with the top 20% of predicted risk are screened in combination including vs excluding clinical routine measure by number of clinical measures in panel. *Absolute differences greater than 1% were reported in the figure, all values are available in Supplementary Table 7. In each box, grey numbers indicate combinations including vs excluding the given clinical measure. Abbreviations: BMI: Body mass index; LDL: low density cholesterol; HDL: high density cholesterol; ALAT: Alanine aminotransferase; ASAT: aspartate aminotransferase; GGT: gamma-glutamyl transferase; eGFR: estimated glomerular filtration rate; HbA1c: glycated hemoglobin A1C.

Sensitivity analyses replicating the main model in subgroups of participants of different ages and in subgroups defined by the presence of metabolic and/or cardiometabolic diseases show consistent results (**Supplementary Table 8, Supplementary Figures 4 and 5**).

## Discussion

Rather than seeking another diagnostic biomarker of frailty that may be expensive to collect and inaccessible to many, we investigated whether routinely collected clinical data could improve the efficiency of an existing clinical screening strategy using data from more than 280,000 adults living with chronic diseases. Three main findings emerged. First, routine clinical measures substantially enriched frailty detection: the 20% of individuals at highest predicted risk accounted for more than half of all frailty cases. Second, the clinical profile of the population prioritized by the model was consistent with the intended use of a frailty screening strategy. Frail individuals always exhibited markedly poorer health profiles regarding mortality, self-rated health, polypharmacy and falls compared to non-frail individuals. Third, performance remained stable across alternative clinical combinations and participant subgroups, suggesting that risk-prioritization did not depend on a specific set of clinical tests or modelling strategy. Taken together, these findings indicate that routinely collected clinical information could provide a pragmatic, scalable “triage” tool to improving the efficiency of frailty screening among adults living with chronic diseases.

Existing frailty guidelines primarily focus on how frailty should be assessed once evaluation is undertaken^16–18^. Little attention has been paid to identifying efficient strategies for selecting which individuals should undergo initial assessment. As a result, recommendations often default to age-based or disease-based eligibility criteria despite limited empirical evidence supporting these approaches. Although chronic diseases are major risk factors for frailty, most individuals living with chronic diseases are not frail. Previous studies have shown that while a large proportion of frail individuals have multimorbidity, only a minority of individuals with multimorbidity are themselves frail^24^. In our study among individuals with at least one chronic condition, frailty prevalence was approximately 6%, consistent with previous reports among middle-aged adults living with chronic diseases^2^ and remaining below 10% across most disease groups in this age range^7^. Consequently, universal frailty assessment within chronic disease populations will identify the individuals with frailty, but at the immense cost of needing to screen everyone, most of whom will not be identified as frail.

Although an increasing number of chronic disease guidelines recommend assessing frailty,^8–10^ they generally provide limited guidance regarding whom to assess and when assessment should begin, leaving chronological age as the default criterion in routine practice. We note that approximately 62% of adults aged 65 years or older in Europe report at least one chronic disease, with this proportion approaching 76% among the oldest adults^31^ and 100% in hospitalized older adults^32^. A screening strategy focusing on individuals living with chronic diseases would therefore naturally include a large share of older adults while extending assessment to younger individuals experiencing accelerated physiological decline. Consistent with this interpretation, the risk-prioritization achieved by our model remained stable across age groups or in subgroups with cardio(metabolic) diseases.

Our findings support an alternative approach in which routinely available clinical information is used to prioritize those most likely to be diagnosed as frail. To our knowledge, no previous large-scale study has formally investigated the efficiency of frailty screening strategies, and current recommendations largely rely on expert consensus regarding whom to assess^15–18^. Whether prediction-based strategy ultimately improves patient outcomes remains to be determined. Because the model is intended only to prioritize clinical assessment, predictions that are false-positive primarily will likely only result in additional frailty assessment rather than inappropriate treatment decisions. We also noted that individuals with high predicted risk who were not classified as frail nevertheless exhibited poorer health profiles, suggesting that this group may nonetheless warrant closer clinical attention although frailty seems not to be involved.

Electronic frailty indices (eFIs) have emerged as promising approaches for the direct identification and monitoring of frailty at scale^33^. eFIs constitutes a different framework for frailty, designed for population-level risk stratification, whereas the Fried phenotype underpins a clinical approach and individual interventions. The overlap between the two approaches of the same construct is moderate^34^. These approaches may not be viewed as competing but rather as complementary tools addressing different clinical and research needs^14^. Importantly, most intervention trials evaluating strategies to prevent or manage frailty have not been designed around frailty indices but have frequently relied on the Fried physical frailty phenotype^11,12^. In the context of routine measurements, eFIs also require a larger set of variables, which may be more complicated to collect.

Routine clinical testing is not standardized across healthcare systems, clinical specialties, or chronic diseases. It reflects a combination of guideline recommendations, local practice patterns, and disease-specific monitoring requirements^35^. We noted that the gain advantage of the clinical-guided approach remained robust despite substantial variation in the clinical panels evaluated, chronic disease definitions, and participant characteristics. No individual clinical measure appeared indispensable, while gain improved progressively as additional clinical information was incorporated. These observations suggest that the stability of the approach arises not from any specific biomarker but from the aggregate physiological information captured by routine clinical testing. In our example, although the strategy assumes that the measures are routinely collected as part of the monitoring of individuals with chronic illnesses, the combination of 12 measures costs approximately 37 euros.

However, we observe shorter combinations with similar performance; cost does not appear to reflect effectiveness. From an implementation perspective, this stability is encouraging because it suggests that clinical-guided frailty prioritization may remain applicable despite the considerable heterogeneity in clinical practice observed across diseases and healthcare systems. Reproducibility may be sought at the level of the screening strategy rather than the exact clinical panel.

Previous work sought clinical biomarkers capable of diagnosing frailty^36^. Most biomarker studies have focused on identifying specific biological signatures, including inflammatory markers, endocrine pathways, or immune biomarkers profiles^37^. Despite promising associations, few have translated into clinical practice, partly because frailty may emerge from multisystem dysregulation rather than a single biological pathway^38^. Moreover, discrimination metrics such as the ROC-AUC provide limited insight into the practical value of models for rare status, where the clinical priority is case identification^39^. Our results suggest that, pending the development of a biomarker capable of reliably and directly identifying physical frailty, a clinical-guided triage tool screening could be a valid approach.

Beyond frailty, these findings illustrate how routinely collected healthcare data could support preventive medicine^40,41^. By repurposing clinical information generated during chronic disease management, frailty prioritization could be embedded within existing care pathways without additional testing or specialized assays. The value of this approach lies not only in identifying individuals at increased risk, but also in potentially facilitating timely access to established interventions, including physical activity programs and nutritional support^11,12^. Rather than creating a new screening program, clinical-guided prioritization may enable existing multidisciplinary services to reach those most likely to benefit.

Several limitations should be considered. First, our model was designed to prioritize individuals for clinical frailty assessment rather than to diagnose frailty directly. Comprehensive clinical evaluation remains essential to confirm frailty, identify contributing factors, and guide personalized interventions. Second, UK Biobank participants are generally healthier than the underlying UK population, and the prevalence of frailty may therefore be underestimated. Although this limitation is unlikely to alter the comparative evaluation of screening strategies, external validation in healthcare systems with different laboratory practices, patient populations and electronic health record infrastructures will be mandatory. Although external validation remains necessary, the robustness observed across heterogeneous clinical panels and disease groups suggests that the underlying signal may be transferable across healthcare settings where routine clinical practice differs substantially. Fourth, the optimal proportion of individuals prioritized for assessment will depend on healthcare capacity, prevalence of frailty, and the resources available for subsequent assessment and intervention.

The principal challenge in frailty screening may now not be in determining how to assess frailty, but identifying whom to assess. In a large population of adults living with chronic diseases, clinical measures that are routinely collected improved the efficiency of identifying individuals most likely to be frail. Whether clinical-guided prioritization improves access to effective frailty interventions and ultimately patients’ outcomes remains to be determined.

## Contribution statement

**Benjamin LANDRÉ**: Conceptualization, methodology, software, formal analysis, data curation, writing original draft and editing, funding acquisition, supervision.; **Gabriella C SILVA**: conceptualization, validation, writing – review & editing; **Aurore FAYOSSE**: software, validation, data curation, writing – review & editing; **Ian Meneghel DANILEVICZ**: conceptualization, validation, writing – review & editing; **Wolfgang LIEB**: validation, writing – review & editing, funding acquisition; **Séverine SABIA**: validation, writing – review & editing, funding acquisition; **Archana SINGH-MANOUX**: validation, writing – review & editing, funding acquisition; **Paula STÜRMER:** conceptualization, validation, writing – review & editing, funding acquisition, supervision.

## Acknowledgment

BL is funded by a grant from “France 2030 ANR-23-PAVH-0006” (https://ppr-autonomie.com/le-ppr-autonomie/) and received a salary. GCS is funded by IReSP/CNSA as part of the 2023 call for projects for the research support program “Older people, people with disabilities at all stages of life, relatives, and professionals,” supported by the National Solidarity Fund for Autonomy (CNSA) “AAP-2023-APAOB-318115” (https://iresp.net/appel_a_projets/appel-a-projets-2023-analyse-des-politiques-de-lautonomie-de-loffre-medico-sociale-et-des-besoins-des-publics-apaob-session-1/) and received a salary. SS and IMD funded by the European Union (ERC grant number 101043884) (https://erc.europa.eu/homepage), and SS also by the Fondation Alzheimer (1143650) (https://www.fondation-alzheimer.org/en/) and the Fondation Vaincre Alzheimer (FR-23063CB) (https://www.vaincrealzheimer.org/), and IMD receive a salary. PS was supported by a scholarship of the German Academic Exchange Service (DAAD).

We would also like to thank the participants in the UK Biobank cohort, as well as everyone involved in the data collection.

## Data Availability

UKB data is available, upon request, by following the application procedure detailed in https://www.ukbiobank.ac.uk/use-our-data/apply-for-access. This work uses data provided by patients and collected by the NHS as part of their care and support.

