## Supplementary materials for "Use of routine clinical data in adults with chronic diseases for identifying risk of frailty: results from the UK Biobank study"

**Supplementary Table 1. List of chronic conditions and corresponding ICD-10 codes used to define multimorbidity indicators**

| **Condition** | **ICD-10 code** | **ICD-10 description (from UK Biobank)** | **UK Biobank Field ID [Source]** | **UK Biobank Field ID [Date/Age]** |
| --- | --- | --- | --- | --- |
| Asthma | J45 | Asthma | 131495 | 131494 |
| Atrial fibrillation | I48  I49 | Atrial fibrillation and flutter  Other cardiac arrhythmias | 131351  131353 | 131350  131352 |
| Bronchiectasis | J47 | Bronchiectasis | 131499 | 131498 |
| Cancer | *This was derived using the age at cancer diagnosis from central registry* | | | 40008 |
| Chronic kidney disease | N17  N18  N19  N00  N01  N03  N04  N05  N08  I12  I13 | Acute renal failure  Chronic renal failure  Unspecified renal failure  Acute nephritic syndrome  Rapidly progressive nephritic syndrome  Chronic nephritic syndrome  Nephrotic syndrome  Unspecified nephritic syndrome  Glomerular disorders in diseases classified elsewhere  Hypertensive renal disease  Hypertensive heart and renal disease | 132031  132033  132035  131999  132001  132005  132007  132009  132015  131291  131293 | 132030  132032  132034  131998  132000  132004  132006  132008  132014  131290  131292 |
| Chronic obstructive pulmonary disorder (COPD) | J44  J43  J42  J41  J40 | Other COPD  Emphysema  Unspecified Chronic Bronchitis  Simple and mucopurulent chronic bronchitis  Bronchitis, not specified as acute or chronic | 131493  131491  131489  131487  131485 | 131492  131490  131488  131486  131484 |
| Chronic sinusitis | J32 | Chronic sinusitis | 131469 | 131468 |
| Connective tissue disorders | K90  M05  M06  M07  M08  M30  M31  M32  M33  M34  M35  M36  M60 | Intestinal malabsorption  Seropositive rheumatoid arthritis  Other rheumatoid arthritis  Psoriatic and enteropathic arthropathies  Juvenile arthritis  Polyarteritis nodosa and related conditions  Other necrotising vasculopathies  Systemic lupus erythematosus  Dermatopolymyositis  Systemic sclerosis  Other systemic involvement of connective tissue  Systemic disorders of connective tissue in diseases classified elsewhere  Myositis | 131689  131849  131851  131853  131855  131891  131893  131895  131897  131899  131901  131903  131931 | 131688  131848  131850  131852  131854  131890  131892  131894  131896  131898  131900  131902  131930 |
| Coronary heart disease | I20  I21  I22  I23  I24  I25 | Angina pectoris  Acute myocardial infarction  Subsequent myocardial infarction  Certain current complications following acute myocardial infarction  Other acute ischaemic heart diseases  Chronic ischaemic heart disease | 131297  131299  131301  131303  131305  131307 | 131296  131298  131300  131302  131304  131306 |
| Dementia | F01  F02  F03  G30  G31 | Vascular dementia  Dementia in other diseases classified elsewhere  Unspecified dementia  Alzheimer’s disease  Other degenerative diseases of nervous system, not elsewhere classified | 130839  130841  130843  131037  131039 | 130838  130840  130842  131036  131038 |
| Diabetes | E11  E10  E13  O24 | Non-insulin-dependent diabetes mellitus  Insulin-dependent diabetes mellitus  Other specified diabetes mellitus  Diabetes mellitus in pregnancy | 130709  130707  130713  132203 | 130708  130706  130712  132202 |
| Diverticular disease | K57 | Diverticular disease of intestine | 131637 | 131636 |
| Dyspepsia/ulcer | K21  K22  K25  K26  K27  K28  K29  K30 | Gastro-oesophageal reflux disease  Other diseases of oesophagus  Gastric ulcer  Duodenal ulcer  Peptic ulcer, site unspecified  Gastrojejunal ulcer  Gastritis and duodenitis  Dyspepsia | 131585  131587  131591  131593  131595  131597  131599  131601 | 131584  131586  131590  131592  131594  131596  131598  131600 |
| Endometriosis | N80 | Endometriosis | 132123 | 132122 |
| Epilepsy | G40 | Epilepsy | 131049 | 131048 |
| Glaucoma | H40  H42 | Glaucoma  Glaucoma in diseases classified elsewhere | 131187  131189 | 131186  131188 |
| Heart failure | I46  I50 | Cardiac arrest  Heart failure | 131347  131355 | 131346  131354 |
| Hepatitis | B15  B16  B17  B18  B19 | Acute Hepatitis A  Acute Hepatitis B  Other acute viral hepatitis  Chronic viral hepatitis  Unspecified viral hepatitis | 130195  130197  130199  130201  130203 | 130194  130196  130198  130200  130202 |
| Hypertension | I10  I11  I12  I13  I15 | Essential (primary) hypertension  Hypertensive heart disease  Hypertensive renal disease  Hypertensive heart and renal disease  Secondary hypertension | 131287  131289  131291  131293  131295 | 131286  131288  131290  131292  131294 |
| Inflammatory bowel disease | K50  K51  K52 | Crohn’s disease [regional enteritis]  Ulcerative colitis  Other non-infective gastro-enteritis and colitis | 131627  131629  131631 | 131626  131628  131630 |
| Irritable bowel syndrome | K58 | Irritable bowel syndrome | 131639 | 131638 |
| Liver disease | K70  K71  K72  K73  K74  K75  K76  K77 | Alcoholic liver disease  Toxic liver disease  Hepatic failure, not elsewhere classified  Chronic hepatitis, not elsewhere classified  Fibrosis and cirrhosis of liver  Other inflammatory liver diseases  Other diseases of liver  Liver disorders in diseases classified elsewhere | 131659  131661  131663  131665  131667  131669  131671  131673 | 131658  131660  131662  131664  131666  131668  131670  131672 |
| Migraine | G43 | Migraine | 131053 | 131052 |
| Multiple sclerosis | G35 | Multiple sclerosis | 131043 | 131042 |
| Osteoporosis | M80  M81  M82 | Osteoporosis with pathological fracture  Osteoporosis without pathological fracture  Osteoporosis in diseases classified elsewhere | 131963  131965  131967 | 131962  131964  131966 |
| Painful conditions | M45  M46  M47  M48  M49  M50  M51  M53  M54  G44  M10  M11  M12  M13  M14  M79  B02 | Ankylosing spondylitis  Other inflammatory spondylopathies  Spondylosis  Other spondylopathies  Spondylopathies in diseases classified elsewhere  Cervical disk disorders  Other intervertebral disk disorders  Other dorsopathies, not elsewhere classified  Dorsalgia  Other headache syndromes  Gout  Other crystal arthropathies  Other specific arthropathies  Other arthritis  Arthropathies in other diseases classified elsewhere  Other soft tissue diseases, not elsewhere classified  Zoster [herpes zoster] | 131913  131915  131917  131919  131921  131923  131925  131927  131929  131055  131859  131861  131863  131865  131867  131961  130179 | 131912  131914  131916  131918  131920  131922  131924  131926  131928  131054  131858  131860  131862  131864  131866  131960  130178 |
| Parkinson’s disease | G20  G21 | Parkinson’s disease  Secondary parkinsonism | 131023  131025 | 131022  131024 |
| Peripheral vascular disease | I70  I73 | Atherosclerosis  Other peripheral vascular diseases | 131381  131387 | 131380  131386 |
| Pernicious anaemia | D51 | Vitamin B12 Deficiency Anaemia | 130625 | 130624 |
| Prostate conditions (not cancer) | N40  N41  N42 | Hyperplasia of prostate  Inflammatory diseases of prostate  Other disorders of prostate | 132073  132075  132077 | 132072  132074  132076 |
| Psoriasis/eczema | L20  L21  L22  L23  L24  L25  L26  L27  L28  L29  L30  L40  L41 | Atopic dermatitis  Seborrhoeic dermatitis  Diaper [napkin] dermatitis  Allergic contact dermatitis  Irritant contact dermatitis  Unspecified contact dermatitis  Exfoliative dermatitis  Dermatitis due to substances taken internally  Lichen simplex chronicus and prurigo  Pruritus  Other dermatitis  Psoriasis  Parapsoriasis | 131721  131723  131725  131727  131729  131731  131733  131735  131737  131739  131741  131743  131745 | 131720  131722  131724  131726  131728  131730  131732  131734  131736  131738  131740  131742  131744 |
| Stroke/transient ischaemic attack | I60  I61  I62  I63  I65  I66  I67  I68  I69 | Subarachnoid haemorrhage  Intracerebral haemorrhage  Other nontraumatic intracranial haemorrhage  Cerebral infarction  Occlusion and stenosis of precerebral arteries, not resulting in cerebral infarction  Occlusion and stenosis of cerebral arteries, not resulting in cerebral infarction  Other cerebrovascular diseases  Cerebrovascular disorders in diseases classified elsewhere  Sequelae of cerebrovascular disease | 131361  131363  131365  131367  131371  131373  131375  131377  131379 | 131360  131362  131364  131366  131370  131372  131374  131376  131378 |
| Thyroid conditions | E00  E01  E02  E03  E04  E05  E06  E07 | Congenital iodine-deficiency syndrome  Iodine-deficiency-related thyroid disorders and allied conditions  Subclinical iodine-deficiency hypothyroidism  Other hypothyroidism  Other non-toxic goitre  Thyrotoxicosis [hyperthyroidism]  Thyroiditis  Other disorders of thyroid | 130691  130693  130695  130697  130699  130701  130703  130705 | 130690  130692  130694  130696  130698  130700  130702  130704 |
| Depression | F32  F33  F34  F38  F39 | Depressive episode  Recurrent depressive disorder  Persistent mood [affective] disorders  Other mood [affective] disorders  Unspecified mood [affective] disorder | 130895  130897  130899  130901  130903 | 130894  130896  130898  130900  130902 |
| Anxiety | F41  F42  F43 | Other anxiety disorders  Obsessive-compulsive disorder  Reaction to severe stress, and adjustment disorders | 130907  130909  130911 | 130906  130908  130910 |
| Alcohol/substance dependency | F10  F11  F12  F13  F14  F15  F16  F19 | Mental and behavioral disorders due to use of alcohol  Mental and behavioral disorders due to use of opioids  Mental and behavioral disorders due to use of cannabinoids  Mental and behavioral disorders due to use of sedatives or hypnotics  Mental and behavioral disorders due to use of cocaine  Mental and behavioral disorders due to use of other stimulants, incl. caffeine  Mental and behavioral disorders due to use of hallucinogens  Mental and behavioral disorders due to use of multiple drug use and use of other psychoactive substances | 130855  130857  130859  130861  130863  130865  130867  130873 | 130854  130856  130858  130860  130862  130864  130866  130872 |
| Severe mental illness | F20  F21  F22  F23  F24  F25  F28  F29  F30  F31 | Schizophrenia  Schizotypal disorder  Persistent delusional disorders  Acute and transient psychotic disorders  Induced delusional disorder  Schizoaffective disorders  Other nonorganic psychotic disorders  Unspecified nonorganic psychosis  Manic episode  Bipolar affective disorder | 130875  130877  130879  130881  130883  130885  130887  130889  130891  130893 | 130874  130876  130878  130880  130882  130884  130886  130888  130890  130892 |
| Anorexia or bulimia | F50 | Eating disorders | 130919 | 130918 |

ICD-10 codes for chronic conditions were based on the list of codes used to define these conditions in Ronaldson et al.

**Supplementary Table 2. Listing of the 821 combinations of clinical measures**

| **Panel** | **Panel size** | **BMI** | **Total cholesterol** | **LDL** | **HDL** | **Triglycerides** | **ALAT** | **ASAT** | **GGT** | **Creatinine** | **eGFR** | **HbA1c** | **Glucose** |
| --- | --- | --- | --- | --- | --- | --- | --- | --- | --- | --- | --- | --- | --- |
| P001 | 3 |  |  |  | ✓ |  |  |  |  | ✓ | ✓ |  |  |
| P002 | 3 |  |  |  |  |  |  | ✓ |  | ✓ |  |  | ✓ |
| P003 | 3 |  |  |  |  | ✓ |  |  | ✓ |  |  |  | ✓ |
| P004 | 3 | ✓ |  | ✓ |  |  |  | ✓ |  |  |  |  |  |
| P005 | 3 |  |  |  |  |  | ✓ |  |  | ✓ | ✓ |  |  |
| P006 | 3 |  |  |  |  | ✓ | ✓ |  |  |  |  |  | ✓ |
| P007 | 3 | ✓ |  |  |  |  |  |  |  | ✓ | ✓ |  |  |
| P008 | 3 |  |  | ✓ |  |  | ✓ |  |  | ✓ |  |  |  |
| P009 | 3 | ✓ |  |  |  |  |  | ✓ |  |  | ✓ |  |  |
| P010 | 3 |  |  |  |  |  |  |  |  | ✓ | ✓ | ✓ |  |
| P011 | 3 |  |  |  |  |  |  |  | ✓ | ✓ |  |  | ✓ |
| P012 | 3 |  |  |  | ✓ |  |  | ✓ |  |  |  | ✓ |  |
| P013 | 3 |  | ✓ |  |  |  |  | ✓ |  |  |  |  | ✓ |
| P014 | 3 |  | ✓ |  |  |  |  |  | ✓ | ✓ |  |  |  |
| P015 | 3 |  |  |  |  |  | ✓ |  |  | ✓ |  |  | ✓ |
| P016 | 3 |  |  |  |  |  |  |  |  | ✓ |  | ✓ | ✓ |
| P017 | 3 |  |  |  |  | ✓ |  |  |  |  |  | ✓ | ✓ |
| P018 | 3 |  | ✓ |  |  |  |  |  | ✓ |  | ✓ |  |  |
| P019 | 3 |  |  |  | ✓ | ✓ | ✓ |  |  |  |  |  |  |
| P020 | 3 |  | ✓ |  |  |  |  |  |  |  | ✓ |  | ✓ |
| P021 | 3 |  | ✓ |  | ✓ |  |  |  |  |  |  |  | ✓ |
| P022 | 3 | ✓ |  |  | ✓ |  |  |  |  |  |  | ✓ |  |
| P023 | 3 | ✓ | ✓ |  |  |  |  |  |  | ✓ |  |  |  |
| P024 | 3 |  |  |  |  |  |  |  | ✓ |  | ✓ |  | ✓ |
| P025 | 3 |  |  |  |  |  |  | ✓ | ✓ |  | ✓ |  |  |
| P026 | 3 |  |  |  | ✓ |  |  |  |  |  |  | ✓ | ✓ |
| P027 | 3 |  | ✓ |  |  | ✓ |  |  |  |  |  | ✓ |  |
| P028 | 3 |  | ✓ |  |  |  | ✓ |  | ✓ |  |  |  |  |
| P029 | 3 |  |  |  |  |  |  |  | ✓ | ✓ |  | ✓ |  |
| P030 | 3 |  |  | ✓ | ✓ |  |  | ✓ |  |  |  |  |  |
| P031 | 3 |  |  | ✓ |  |  | ✓ |  | ✓ |  |  |  |  |
| P032 | 3 |  | ✓ |  |  | ✓ |  |  |  | ✓ |  |  |  |
| P033 | 3 |  |  |  | ✓ | ✓ |  |  |  |  |  |  | ✓ |
| P034 | 3 | ✓ |  |  |  | ✓ |  |  |  |  | ✓ |  |  |
| P035 | 3 |  |  | ✓ |  | ✓ | ✓ |  |  |  |  |  |  |
| P036 | 3 |  |  |  |  |  | ✓ |  |  |  | ✓ | ✓ |  |
| P037 | 3 |  |  |  |  |  | ✓ |  |  | ✓ |  | ✓ |  |
| P038 | 3 |  |  |  |  | ✓ | ✓ |  |  |  |  | ✓ |  |
| P039 | 3 |  | ✓ |  |  | ✓ |  | ✓ |  |  |  |  |  |
| P040 | 3 | ✓ |  |  | ✓ |  |  |  | ✓ |  |  |  |  |
| P041 | 3 |  |  |  | ✓ |  |  |  | ✓ | ✓ |  |  |  |
| P042 | 3 | ✓ |  |  |  |  |  |  |  |  | ✓ | ✓ |  |
| P043 | 3 |  |  | ✓ |  |  |  |  |  |  | ✓ |  | ✓ |
| P044 | 3 |  |  |  |  |  |  |  |  | ✓ | ✓ |  | ✓ |
| P045 | 4 |  |  |  | ✓ |  |  | ✓ |  | ✓ |  | ✓ |  |
| P046 | 4 |  | ✓ |  | ✓ |  |  |  |  | ✓ | ✓ |  |  |
| P047 | 4 |  | ✓ | ✓ | ✓ | ✓ |  |  |  |  |  |  |  |
| P048 | 4 |  | ✓ |  | ✓ |  |  | ✓ |  |  | ✓ |  |  |
| P049 | 4 |  |  | ✓ | ✓ | ✓ |  |  |  |  | ✓ |  |  |
| P050 | 4 |  |  |  |  |  | ✓ |  |  | ✓ | ✓ | ✓ |  |
| P051 | 4 | ✓ |  | ✓ |  |  |  | ✓ |  |  | ✓ |  |  |
| P052 | 4 | ✓ |  | ✓ |  |  |  |  | ✓ | ✓ |  |  |  |
| P053 | 4 | ✓ |  | ✓ |  |  |  |  |  | ✓ | ✓ |  |  |
| P054 | 4 | ✓ |  | ✓ |  |  | ✓ |  |  | ✓ |  |  |  |
| P055 | 4 | ✓ |  |  |  |  | ✓ |  |  | ✓ |  | ✓ |  |
| P056 | 4 |  | ✓ |  | ✓ |  | ✓ |  | ✓ |  |  |  |  |
| P057 | 4 |  |  | ✓ |  |  |  | ✓ | ✓ |  |  |  | ✓ |
| P058 | 4 |  |  | ✓ |  |  | ✓ |  |  |  | ✓ | ✓ |  |
| P059 | 4 | ✓ |  |  |  |  |  | ✓ |  |  | ✓ | ✓ |  |
| P060 | 4 |  |  | ✓ | ✓ |  | ✓ |  | ✓ |  |  |  |  |
| P061 | 4 |  | ✓ |  |  |  |  |  | ✓ | ✓ |  | ✓ |  |
| P062 | 4 |  |  |  |  |  | ✓ | ✓ | ✓ |  |  | ✓ |  |
| P063 | 4 | ✓ | ✓ |  |  |  |  |  |  | ✓ |  | ✓ |  |
| P064 | 4 |  |  |  |  | ✓ | ✓ |  | ✓ | ✓ |  |  |  |
| P065 | 4 | ✓ |  |  | ✓ |  | ✓ |  | ✓ |  |  |  |  |
| P066 | 4 |  |  | ✓ |  | ✓ | ✓ |  |  | ✓ |  |  |  |
| P067 | 4 |  | ✓ |  | ✓ |  |  |  | ✓ |  |  |  | ✓ |
| P068 | 4 | ✓ | ✓ |  | ✓ |  |  |  |  |  |  | ✓ |  |
| P069 | 4 | ✓ |  |  |  | ✓ |  | ✓ | ✓ |  |  |  |  |
| P070 | 4 | ✓ |  |  | ✓ |  | ✓ |  |  |  |  |  | ✓ |
| P071 | 4 |  | ✓ |  |  |  | ✓ |  |  | ✓ |  |  | ✓ |
| P072 | 4 |  | ✓ |  |  | ✓ | ✓ |  |  |  |  |  | ✓ |
| P073 | 4 |  |  |  |  | ✓ |  |  | ✓ |  |  | ✓ | ✓ |
| P074 | 4 | ✓ |  |  | ✓ | ✓ |  |  |  |  | ✓ |  |  |
| P075 | 4 |  |  | ✓ |  |  | ✓ |  | ✓ |  |  | ✓ |  |
| P076 | 4 | ✓ | ✓ |  |  |  |  |  | ✓ |  |  |  | ✓ |
| P077 | 4 | ✓ |  |  |  |  |  |  | ✓ |  | ✓ | ✓ |  |
| P078 | 4 |  | ✓ |  |  | ✓ |  | ✓ |  |  |  |  | ✓ |
| P079 | 4 |  | ✓ |  | ✓ |  | ✓ | ✓ |  |  |  |  |  |
| P080 | 4 |  |  |  | ✓ | ✓ | ✓ |  |  |  |  | ✓ |  |
| P081 | 4 |  |  | ✓ | ✓ |  |  |  | ✓ |  |  | ✓ |  |
| P082 | 4 | ✓ | ✓ |  |  |  |  | ✓ |  |  |  | ✓ |  |
| P083 | 4 | ✓ | ✓ | ✓ |  |  |  | ✓ |  |  |  |  |  |
| P084 | 4 | ✓ | ✓ |  | ✓ |  |  |  | ✓ |  |  |  |  |
| P085 | 4 |  |  |  |  |  |  | ✓ |  | ✓ |  | ✓ | ✓ |
| P086 | 4 |  |  |  | ✓ | ✓ |  |  | ✓ |  |  | ✓ |  |
| P087 | 4 |  | ✓ |  |  | ✓ |  |  | ✓ |  |  | ✓ |  |
| P088 | 4 |  |  |  | ✓ |  |  | ✓ | ✓ |  |  |  | ✓ |
| P089 | 4 |  |  | ✓ | ✓ |  |  |  |  | ✓ | ✓ |  |  |
| P090 | 4 |  | ✓ |  |  |  |  |  | ✓ | ✓ |  |  | ✓ |
| P091 | 4 | ✓ |  |  | ✓ |  | ✓ | ✓ |  |  |  |  |  |
| P092 | 4 |  |  |  | ✓ |  |  |  | ✓ | ✓ | ✓ |  |  |
| P093 | 4 | ✓ | ✓ |  |  |  | ✓ | ✓ |  |  |  |  |  |
| P094 | 4 |  |  | ✓ | ✓ | ✓ |  |  |  |  |  | ✓ |  |
| P095 | 4 |  |  |  | ✓ |  |  |  |  | ✓ |  | ✓ | ✓ |
| P096 | 4 |  |  | ✓ |  |  |  |  |  | ✓ |  | ✓ | ✓ |
| P097 | 4 |  |  | ✓ | ✓ | ✓ | ✓ |  |  |  |  |  |  |
| P098 | 4 |  |  |  | ✓ |  | ✓ |  | ✓ | ✓ |  |  |  |
| P099 | 4 |  |  |  |  |  | ✓ |  |  | ✓ | ✓ |  | ✓ |
| P100 | 4 | ✓ |  |  |  | ✓ |  |  | ✓ | ✓ |  |  |  |
| P101 | 4 | ✓ |  |  |  | ✓ | ✓ | ✓ |  |  |  |  |  |
| P102 | 4 | ✓ |  |  |  |  |  |  | ✓ | ✓ |  |  | ✓ |
| P103 | 4 | ✓ |  | ✓ |  |  | ✓ |  |  |  | ✓ |  |  |
| P104 | 4 |  |  |  | ✓ |  | ✓ |  | ✓ |  |  | ✓ |  |
| P105 | 4 |  | ✓ | ✓ |  |  |  |  |  |  | ✓ | ✓ |  |
| P106 | 4 | ✓ |  | ✓ |  |  |  | ✓ | ✓ |  |  |  |  |
| P107 | 4 | ✓ |  |  |  |  |  | ✓ |  | ✓ |  | ✓ |  |
| P108 | 4 |  |  |  | ✓ | ✓ |  | ✓ |  |  | ✓ |  |  |
| P109 | 4 |  |  | ✓ |  |  | ✓ | ✓ | ✓ |  |  |  |  |
| P110 | 4 | ✓ |  |  | ✓ | ✓ |  |  |  | ✓ |  |  |  |
| P111 | 4 | ✓ |  |  |  |  |  |  |  |  | ✓ | ✓ | ✓ |
| P112 | 4 | ✓ |  |  |  |  | ✓ |  | ✓ | ✓ |  |  |  |
| P113 | 4 | ✓ |  | ✓ | ✓ |  |  |  |  |  | ✓ |  |  |
| P114 | 4 | ✓ |  | ✓ |  |  |  |  | ✓ |  |  | ✓ |  |
| P115 | 4 |  | ✓ | ✓ |  | ✓ |  |  |  |  | ✓ |  |  |
| P116 | 4 |  |  | ✓ |  |  |  |  | ✓ | ✓ |  |  | ✓ |
| P117 | 4 |  | ✓ |  |  | ✓ |  | ✓ | ✓ |  |  |  |  |
| P118 | 4 | ✓ |  |  | ✓ |  |  | ✓ |  |  |  | ✓ |  |
| P119 | 4 |  |  | ✓ |  | ✓ |  |  |  | ✓ |  | ✓ |  |
| P120 | 4 |  | ✓ |  | ✓ |  | ✓ |  |  |  |  |  | ✓ |
| P121 | 4 | ✓ |  |  |  | ✓ |  |  |  | ✓ |  |  | ✓ |
| P122 | 4 |  | ✓ |  | ✓ |  | ✓ |  |  |  | ✓ |  |  |
| P123 | 4 |  | ✓ | ✓ |  | ✓ | ✓ |  |  |  |  |  |  |
| P124 | 4 |  | ✓ |  |  |  |  | ✓ |  |  | ✓ | ✓ |  |
| P125 | 4 |  |  | ✓ | ✓ |  |  | ✓ |  |  |  | ✓ |  |
| P126 | 4 |  |  | ✓ | ✓ |  |  |  |  | ✓ |  |  | ✓ |
| P127 | 4 |  | ✓ |  |  | ✓ | ✓ |  |  | ✓ |  |  |  |
| P128 | 4 |  |  | ✓ |  |  |  |  |  | ✓ | ✓ | ✓ |  |
| P129 | 4 |  |  |  |  | ✓ |  |  | ✓ | ✓ |  |  | ✓ |
| P130 | 4 |  |  | ✓ |  |  |  | ✓ | ✓ | ✓ |  |  |  |
| P131 | 4 |  |  |  |  |  | ✓ |  | ✓ |  |  | ✓ | ✓ |
| P132 | 4 |  | ✓ |  |  |  |  |  | ✓ |  | ✓ |  | ✓ |
| P133 | 4 | ✓ |  |  |  | ✓ | ✓ |  |  |  | ✓ |  |  |
| P134 | 4 | ✓ |  |  | ✓ |  |  |  |  |  | ✓ | ✓ |  |
| P135 | 4 |  |  |  | ✓ |  | ✓ | ✓ | ✓ |  |  |  |  |
| P136 | 4 |  |  |  |  | ✓ | ✓ |  | ✓ |  |  | ✓ |  |
| P137 | 4 | ✓ |  |  |  |  |  | ✓ |  |  | ✓ |  | ✓ |
| P138 | 4 | ✓ |  |  | ✓ |  |  |  | ✓ |  |  | ✓ |  |
| P139 | 4 |  | ✓ |  |  |  | ✓ | ✓ |  |  |  |  | ✓ |
| P140 | 4 | ✓ |  |  |  |  |  |  | ✓ |  | ✓ |  | ✓ |
| P141 | 4 |  |  |  | ✓ |  | ✓ |  |  |  |  | ✓ | ✓ |
| P142 | 4 | ✓ |  |  |  |  |  | ✓ |  |  |  | ✓ | ✓ |
| P143 | 4 | ✓ | ✓ | ✓ |  |  |  |  | ✓ |  |  |  |  |
| P144 | 5 | ✓ |  |  |  |  |  |  | ✓ | ✓ | ✓ | ✓ |  |
| P145 | 5 |  |  |  |  |  | ✓ |  | ✓ | ✓ |  | ✓ | ✓ |
| P146 | 5 | ✓ |  |  |  | ✓ |  | ✓ |  | ✓ | ✓ |  |  |
| P147 | 5 |  |  |  |  |  |  | ✓ | ✓ | ✓ |  | ✓ | ✓ |
| P148 | 5 |  |  | ✓ | ✓ |  | ✓ |  | ✓ | ✓ |  |  |  |
| P149 | 5 | ✓ |  |  | ✓ |  | ✓ |  |  |  | ✓ | ✓ |  |
| P150 | 5 |  |  |  |  | ✓ |  | ✓ |  | ✓ | ✓ |  | ✓ |
| P151 | 5 |  | ✓ | ✓ | ✓ |  | ✓ |  | ✓ |  |  |  |  |
| P152 | 5 | ✓ | ✓ |  | ✓ | ✓ |  |  | ✓ |  |  |  |  |
| P153 | 5 | ✓ |  | ✓ | ✓ |  |  | ✓ |  |  |  | ✓ |  |
| P154 | 5 |  | ✓ |  | ✓ |  |  | ✓ |  | ✓ | ✓ |  |  |
| P155 | 5 |  |  |  |  | ✓ | ✓ | ✓ | ✓ |  | ✓ |  |  |
| P156 | 5 |  |  | ✓ | ✓ | ✓ |  |  |  |  | ✓ |  | ✓ |
| P157 | 5 |  |  | ✓ | ✓ |  |  |  | ✓ | ✓ |  |  | ✓ |
| P158 | 5 | ✓ | ✓ |  |  | ✓ |  |  |  |  | ✓ | ✓ |  |
| P159 | 5 |  |  |  | ✓ | ✓ |  |  | ✓ |  | ✓ |  | ✓ |
| P160 | 5 |  |  |  |  |  |  | ✓ |  | ✓ | ✓ | ✓ | ✓ |
| P161 | 5 | ✓ |  | ✓ |  |  |  |  | ✓ | ✓ |  | ✓ |  |
| P162 | 5 |  |  |  |  | ✓ |  |  | ✓ | ✓ |  | ✓ | ✓ |
| P163 | 5 |  |  |  | ✓ | ✓ | ✓ |  |  |  | ✓ |  | ✓ |
| P164 | 5 | ✓ |  |  |  | ✓ |  |  | ✓ | ✓ | ✓ |  |  |
| P165 | 5 |  |  | ✓ |  | ✓ | ✓ |  |  | ✓ | ✓ |  |  |
| P166 | 5 |  | ✓ |  |  | ✓ |  |  | ✓ |  | ✓ |  | ✓ |
| P167 | 5 |  | ✓ | ✓ | ✓ |  |  | ✓ | ✓ |  |  |  |  |
| P168 | 5 | ✓ |  |  |  |  | ✓ |  | ✓ |  | ✓ |  | ✓ |
| P169 | 5 |  | ✓ |  | ✓ |  |  | ✓ |  |  | ✓ |  | ✓ |
| P170 | 5 | ✓ | ✓ | ✓ |  |  | ✓ |  |  |  |  | ✓ |  |
| P171 | 5 | ✓ |  | ✓ |  |  |  |  | ✓ | ✓ | ✓ |  |  |
| P172 | 5 | ✓ |  | ✓ |  | ✓ |  |  |  | ✓ | ✓ |  |  |
| P173 | 5 | ✓ | ✓ |  | ✓ |  |  | ✓ |  |  | ✓ |  |  |
| P174 | 5 |  | ✓ |  |  |  |  | ✓ |  | ✓ |  | ✓ | ✓ |
| P175 | 5 | ✓ |  | ✓ |  |  | ✓ |  | ✓ |  |  | ✓ |  |
| P176 | 5 |  |  | ✓ | ✓ | ✓ |  |  | ✓ |  |  | ✓ |  |
| P177 | 5 | ✓ | ✓ |  |  | ✓ |  |  |  |  | ✓ |  | ✓ |
| P178 | 5 |  | ✓ |  |  |  | ✓ |  |  | ✓ | ✓ |  | ✓ |
| P179 | 5 |  |  | ✓ |  |  | ✓ | ✓ | ✓ |  | ✓ |  |  |
| P180 | 5 |  | ✓ | ✓ |  |  | ✓ |  |  |  | ✓ | ✓ |  |
| P181 | 5 | ✓ |  |  |  |  | ✓ | ✓ |  | ✓ |  |  | ✓ |
| P182 | 5 |  |  | ✓ |  | ✓ | ✓ | ✓ | ✓ |  |  |  |  |
| P183 | 5 |  |  |  | ✓ |  | ✓ | ✓ |  | ✓ | ✓ |  |  |
| P184 | 5 |  |  | ✓ |  | ✓ |  | ✓ |  |  | ✓ | ✓ |  |
| P185 | 5 |  |  |  |  |  | ✓ | ✓ |  | ✓ | ✓ |  | ✓ |
| P186 | 5 |  | ✓ |  | ✓ | ✓ |  |  |  | ✓ | ✓ |  |  |
| P187 | 5 |  |  |  | ✓ |  | ✓ | ✓ |  |  | ✓ |  | ✓ |
| P188 | 5 |  |  |  | ✓ |  | ✓ |  | ✓ | ✓ | ✓ |  |  |
| P189 | 5 |  | ✓ |  | ✓ | ✓ |  |  | ✓ |  |  | ✓ |  |
| P190 | 5 |  |  |  | ✓ | ✓ | ✓ |  | ✓ | ✓ |  |  |  |
| P191 | 5 | ✓ |  |  | ✓ |  |  | ✓ |  |  |  | ✓ | ✓ |
| P192 | 5 |  | ✓ |  | ✓ | ✓ |  |  | ✓ |  |  |  | ✓ |
| P193 | 5 |  | ✓ | ✓ |  |  |  | ✓ | ✓ |  |  |  | ✓ |
| P194 | 5 |  | ✓ |  |  |  |  | ✓ | ✓ | ✓ |  |  | ✓ |
| P195 | 5 |  | ✓ | ✓ |  |  | ✓ | ✓ |  | ✓ |  |  |  |
| P196 | 5 |  |  | ✓ | ✓ | ✓ | ✓ |  |  |  |  | ✓ |  |
| P197 | 5 | ✓ | ✓ |  | ✓ | ✓ |  |  |  | ✓ |  |  |  |
| P198 | 5 |  | ✓ |  |  |  | ✓ |  |  | ✓ | ✓ | ✓ |  |
| P199 | 5 |  | ✓ |  |  | ✓ | ✓ | ✓ |  |  | ✓ |  |  |
| P200 | 5 | ✓ |  | ✓ |  |  |  |  | ✓ |  |  | ✓ | ✓ |
| P201 | 5 | ✓ |  | ✓ | ✓ | ✓ |  |  |  |  | ✓ |  |  |
| P202 | 5 | ✓ |  |  |  | ✓ | ✓ | ✓ |  |  |  |  | ✓ |
| P203 | 5 | ✓ |  | ✓ |  |  |  | ✓ | ✓ |  | ✓ |  |  |
| P204 | 5 |  |  | ✓ | ✓ |  | ✓ |  |  | ✓ |  |  | ✓ |
| P205 | 5 | ✓ |  |  | ✓ |  |  |  | ✓ | ✓ |  | ✓ |  |
| P206 | 5 |  | ✓ |  | ✓ |  |  | ✓ |  |  | ✓ | ✓ |  |
| P207 | 5 | ✓ |  | ✓ |  | ✓ | ✓ |  |  |  | ✓ |  |  |
| P208 | 5 | ✓ | ✓ |  | ✓ |  |  | ✓ |  |  |  |  | ✓ |
| P209 | 5 |  | ✓ |  |  |  |  |  | ✓ | ✓ |  | ✓ | ✓ |
| P210 | 5 | ✓ |  |  | ✓ |  | ✓ |  |  | ✓ | ✓ |  |  |
| P211 | 5 | ✓ |  |  |  | ✓ |  |  | ✓ |  | ✓ | ✓ |  |
| P212 | 5 | ✓ |  | ✓ |  |  |  | ✓ | ✓ | ✓ |  |  |  |
| P213 | 5 |  | ✓ | ✓ |  |  |  |  |  | ✓ |  | ✓ | ✓ |
| P214 | 5 |  |  | ✓ |  | ✓ |  |  | ✓ |  |  | ✓ | ✓ |
| P215 | 5 |  |  |  |  |  | ✓ | ✓ |  |  | ✓ | ✓ | ✓ |
| P216 | 5 | ✓ |  |  |  |  | ✓ |  | ✓ |  | ✓ | ✓ |  |
| P217 | 5 |  |  |  | ✓ |  |  | ✓ |  | ✓ | ✓ | ✓ |  |
| P218 | 5 | ✓ |  |  | ✓ | ✓ | ✓ | ✓ |  |  |  |  |  |
| P219 | 5 |  | ✓ |  |  |  |  |  | ✓ | ✓ | ✓ |  | ✓ |
| P220 | 5 |  |  | ✓ | ✓ |  | ✓ | ✓ |  |  | ✓ |  |  |
| P221 | 5 |  | ✓ | ✓ |  |  | ✓ |  |  | ✓ |  |  | ✓ |
| P222 | 5 |  | ✓ | ✓ |  |  |  |  | ✓ |  | ✓ |  | ✓ |
| P223 | 5 |  | ✓ | ✓ | ✓ |  |  | ✓ |  |  | ✓ |  |  |
| P224 | 5 | ✓ |  | ✓ |  | ✓ |  |  | ✓ | ✓ |  |  |  |
| P225 | 5 |  | ✓ |  | ✓ |  |  |  |  | ✓ | ✓ |  | ✓ |
| P226 | 5 |  | ✓ |  |  |  | ✓ | ✓ | ✓ |  |  |  | ✓ |
| P227 | 5 | ✓ | ✓ |  | ✓ |  |  |  | ✓ |  |  | ✓ |  |
| P228 | 5 |  | ✓ |  | ✓ |  |  | ✓ |  | ✓ |  |  | ✓ |
| P229 | 5 |  |  | ✓ |  | ✓ |  | ✓ |  | ✓ |  | ✓ |  |
| P230 | 5 |  | ✓ | ✓ | ✓ |  |  |  |  |  | ✓ |  | ✓ |
| P231 | 5 | ✓ |  |  |  | ✓ |  | ✓ | ✓ |  |  |  | ✓ |
| P232 | 5 | ✓ |  |  |  | ✓ | ✓ |  |  | ✓ | ✓ |  |  |
| P233 | 5 |  |  | ✓ |  |  | ✓ |  | ✓ |  | ✓ |  | ✓ |
| P234 | 5 |  | ✓ | ✓ | ✓ |  |  | ✓ |  |  |  | ✓ |  |
| P235 | 5 | ✓ |  | ✓ | ✓ |  | ✓ |  | ✓ |  |  |  |  |
| P236 | 5 | ✓ |  |  | ✓ | ✓ |  |  | ✓ |  |  | ✓ |  |
| P237 | 5 |  |  | ✓ |  | ✓ |  | ✓ |  | ✓ |  |  | ✓ |
| P238 | 5 |  |  |  | ✓ | ✓ |  |  |  | ✓ | ✓ | ✓ |  |
| P239 | 5 |  | ✓ | ✓ | ✓ | ✓ |  |  |  |  |  |  | ✓ |
| P240 | 5 | ✓ | ✓ | ✓ |  |  |  | ✓ |  |  |  |  | ✓ |
| P241 | 5 |  | ✓ |  |  |  |  |  | ✓ |  | ✓ | ✓ | ✓ |
| P242 | 5 |  | ✓ |  |  |  | ✓ |  | ✓ |  | ✓ | ✓ |  |
| P243 | 5 |  |  | ✓ |  |  |  |  |  | ✓ | ✓ | ✓ | ✓ |
| P244 | 5 |  |  | ✓ | ✓ | ✓ |  |  | ✓ |  | ✓ |  |  |
| P245 | 5 |  | ✓ | ✓ |  |  | ✓ |  |  | ✓ |  | ✓ |  |
| P246 | 5 |  | ✓ |  |  | ✓ |  |  | ✓ | ✓ |  |  | ✓ |
| P247 | 5 | ✓ |  |  | ✓ | ✓ |  |  |  | ✓ |  |  | ✓ |
| P248 | 5 |  |  |  | ✓ | ✓ | ✓ | ✓ |  |  |  |  | ✓ |
| P249 | 5 |  |  | ✓ |  |  |  | ✓ | ✓ |  |  | ✓ | ✓ |
| P250 | 5 |  | ✓ |  | ✓ | ✓ |  | ✓ | ✓ |  |  |  |  |
| P251 | 5 |  |  |  | ✓ |  | ✓ | ✓ |  |  | ✓ | ✓ |  |
| P252 | 5 |  |  | ✓ |  |  |  | ✓ |  | ✓ |  | ✓ | ✓ |
| P253 | 5 | ✓ |  | ✓ |  | ✓ |  |  | ✓ |  |  |  | ✓ |
| P254 | 5 |  |  |  |  |  |  | ✓ | ✓ | ✓ | ✓ | ✓ |  |
| P255 | 5 |  |  |  | ✓ | ✓ | ✓ |  | ✓ |  | ✓ |  |  |
| P256 | 5 |  |  | ✓ | ✓ |  |  | ✓ | ✓ |  | ✓ |  |  |
| P257 | 5 |  | ✓ |  |  |  | ✓ |  | ✓ | ✓ | ✓ |  |  |
| P258 | 5 | ✓ |  |  |  |  |  |  |  | ✓ | ✓ | ✓ | ✓ |
| P259 | 5 | ✓ |  |  | ✓ | ✓ |  |  |  |  |  | ✓ | ✓ |
| P260 | 5 |  | ✓ | ✓ |  |  | ✓ |  |  | ✓ | ✓ |  |  |
| P261 | 5 | ✓ | ✓ |  |  |  |  |  |  | ✓ | ✓ | ✓ |  |
| P262 | 5 |  |  |  | ✓ |  | ✓ |  | ✓ |  |  | ✓ | ✓ |
| P263 | 5 |  |  | ✓ |  |  | ✓ |  |  | ✓ | ✓ | ✓ |  |
| P264 | 5 | ✓ | ✓ |  | ✓ |  |  |  | ✓ | ✓ |  |  |  |
| P265 | 5 | ✓ |  |  | ✓ | ✓ |  |  | ✓ |  | ✓ |  |  |
| P266 | 5 |  |  |  |  | ✓ | ✓ |  |  | ✓ | ✓ |  | ✓ |
| P267 | 5 |  |  | ✓ | ✓ | ✓ |  |  |  | ✓ |  | ✓ |  |
| P268 | 5 |  |  | ✓ |  |  |  | ✓ |  | ✓ | ✓ | ✓ |  |
| P269 | 5 |  | ✓ |  | ✓ | ✓ | ✓ | ✓ |  |  |  |  |  |
| P270 | 5 | ✓ |  | ✓ | ✓ |  |  | ✓ | ✓ |  |  |  |  |
| P271 | 5 |  | ✓ |  | ✓ |  | ✓ |  |  | ✓ |  |  | ✓ |
| P272 | 5 | ✓ | ✓ |  |  |  |  | ✓ | ✓ |  |  |  | ✓ |
| P273 | 5 |  |  |  | ✓ |  | ✓ | ✓ |  |  |  | ✓ | ✓ |
| P274 | 5 | ✓ |  |  | ✓ | ✓ | ✓ |  |  |  |  |  | ✓ |
| P275 | 5 |  | ✓ | ✓ | ✓ |  |  |  | ✓ | ✓ |  |  |  |
| P276 | 5 |  | ✓ | ✓ |  |  |  | ✓ |  | ✓ |  |  | ✓ |
| P277 | 5 | ✓ |  |  | ✓ |  |  |  |  | ✓ | ✓ |  | ✓ |
| P278 | 5 |  |  |  |  | ✓ |  | ✓ | ✓ |  | ✓ |  | ✓ |
| P279 | 5 |  | ✓ | ✓ |  |  | ✓ |  | ✓ |  | ✓ |  |  |
| P280 | 5 | ✓ |  | ✓ | ✓ |  |  |  | ✓ |  |  | ✓ |  |
| P281 | 5 | ✓ | ✓ | ✓ |  |  |  | ✓ |  |  | ✓ |  |  |
| P282 | 5 |  | ✓ |  | ✓ |  |  |  | ✓ |  |  | ✓ | ✓ |
| P283 | 5 | ✓ |  | ✓ | ✓ |  | ✓ |  |  | ✓ |  |  |  |
| P284 | 5 | ✓ |  | ✓ |  | ✓ |  |  |  | ✓ |  | ✓ |  |
| P285 | 5 |  |  | ✓ | ✓ |  |  |  | ✓ | ✓ |  | ✓ |  |
| P286 | 5 |  | ✓ | ✓ |  | ✓ |  |  |  |  | ✓ | ✓ |  |
| P287 | 5 | ✓ |  | ✓ |  |  | ✓ | ✓ | ✓ |  |  |  |  |
| P288 | 5 |  | ✓ |  | ✓ |  | ✓ |  |  | ✓ | ✓ |  |  |
| P289 | 5 | ✓ |  |  | ✓ |  | ✓ |  | ✓ |  |  |  | ✓ |
| P290 | 5 |  | ✓ |  | ✓ | ✓ |  | ✓ |  | ✓ |  |  |  |
| P291 | 5 |  | ✓ |  |  |  | ✓ | ✓ | ✓ |  |  | ✓ |  |
| P292 | 5 | ✓ | ✓ |  | ✓ |  |  |  |  |  |  | ✓ | ✓ |
| P293 | 5 | ✓ | ✓ |  |  | ✓ |  |  |  | ✓ | ✓ |  |  |
| P294 | 5 |  | ✓ |  |  | ✓ | ✓ |  |  |  | ✓ | ✓ |  |
| P295 | 5 |  |  | ✓ | ✓ | ✓ |  | ✓ |  | ✓ |  |  |  |
| P296 | 5 |  | ✓ |  |  | ✓ | ✓ | ✓ |  |  |  |  | ✓ |
| P297 | 5 | ✓ |  |  |  | ✓ |  |  | ✓ |  |  | ✓ | ✓ |
| P298 | 5 |  | ✓ | ✓ | ✓ |  | ✓ |  |  |  |  |  | ✓ |
| P299 | 5 | ✓ |  |  |  |  |  | ✓ |  | ✓ | ✓ |  | ✓ |
| P300 | 5 |  | ✓ |  |  | ✓ | ✓ |  | ✓ |  |  |  | ✓ |
| P301 | 5 |  |  |  |  |  | ✓ | ✓ |  | ✓ |  | ✓ | ✓ |
| P302 | 5 |  | ✓ |  | ✓ |  | ✓ |  |  |  |  | ✓ | ✓ |
| P303 | 6 | ✓ |  |  |  | ✓ | ✓ | ✓ |  |  | ✓ |  | ✓ |
| P304 | 6 |  | ✓ | ✓ | ✓ | ✓ | ✓ | ✓ |  |  |  |  |  |
| P305 | 6 | ✓ | ✓ |  |  |  | ✓ | ✓ | ✓ |  |  |  | ✓ |
| P306 | 6 |  | ✓ | ✓ |  | ✓ | ✓ |  | ✓ |  |  | ✓ |  |
| P307 | 6 | ✓ | ✓ |  |  |  | ✓ |  |  |  | ✓ | ✓ | ✓ |
| P308 | 6 |  |  | ✓ |  | ✓ |  |  | ✓ |  | ✓ | ✓ | ✓ |
| P309 | 6 | ✓ | ✓ |  | ✓ |  | ✓ |  |  |  | ✓ | ✓ |  |
| P310 | 6 | ✓ |  | ✓ |  | ✓ |  |  |  | ✓ | ✓ |  | ✓ |
| P311 | 6 | ✓ |  | ✓ | ✓ | ✓ |  |  |  |  | ✓ | ✓ |  |
| P312 | 6 | ✓ |  | ✓ | ✓ |  | ✓ |  |  |  | ✓ | ✓ |  |
| P313 | 6 | ✓ | ✓ | ✓ | ✓ |  |  | ✓ | ✓ |  |  |  |  |
| P314 | 6 | ✓ |  |  | ✓ |  | ✓ | ✓ | ✓ |  |  | ✓ |  |
| P315 | 6 |  | ✓ |  |  | ✓ | ✓ | ✓ |  | ✓ |  |  | ✓ |
| P316 | 6 |  | ✓ |  | ✓ | ✓ | ✓ |  |  |  | ✓ |  | ✓ |
| P317 | 6 |  | ✓ |  | ✓ | ✓ | ✓ |  |  |  |  | ✓ | ✓ |
| P318 | 6 |  |  | ✓ | ✓ |  | ✓ |  |  | ✓ |  | ✓ | ✓ |
| P319 | 6 |  | ✓ |  |  |  |  | ✓ | ✓ | ✓ | ✓ | ✓ |  |
| P320 | 6 | ✓ | ✓ |  | ✓ | ✓ |  | ✓ | ✓ |  |  |  |  |
| P321 | 6 | ✓ |  |  | ✓ | ✓ | ✓ |  |  | ✓ |  |  | ✓ |
| P322 | 6 |  | ✓ |  | ✓ |  |  | ✓ | ✓ |  |  | ✓ | ✓ |
| P323 | 6 | ✓ |  |  | ✓ | ✓ |  | ✓ | ✓ |  |  |  | ✓ |
| P324 | 6 |  |  | ✓ |  |  |  |  | ✓ | ✓ | ✓ | ✓ | ✓ |
| P325 | 6 | ✓ | ✓ | ✓ | ✓ |  |  |  |  |  | ✓ | ✓ |  |
| P326 | 6 |  | ✓ | ✓ |  | ✓ | ✓ | ✓ | ✓ |  |  |  |  |
| P327 | 6 | ✓ |  |  |  | ✓ | ✓ |  |  |  | ✓ | ✓ | ✓ |
| P328 | 6 |  |  |  |  | ✓ | ✓ | ✓ |  | ✓ |  | ✓ | ✓ |
| P329 | 6 |  |  | ✓ | ✓ |  | ✓ |  |  | ✓ | ✓ | ✓ |  |
| P330 | 6 | ✓ |  | ✓ | ✓ | ✓ | ✓ | ✓ |  |  |  |  |  |
| P331 | 6 |  | ✓ |  | ✓ | ✓ | ✓ | ✓ |  | ✓ |  |  |  |
| P332 | 6 |  | ✓ |  | ✓ | ✓ | ✓ | ✓ |  |  |  |  | ✓ |
| P333 | 6 |  | ✓ | ✓ |  |  | ✓ | ✓ | ✓ |  | ✓ |  |  |
| P334 | 6 |  |  |  | ✓ | ✓ |  |  | ✓ | ✓ | ✓ | ✓ |  |
| P335 | 6 | ✓ |  |  | ✓ |  | ✓ | ✓ | ✓ |  | ✓ |  |  |
| P336 | 6 |  |  |  | ✓ | ✓ | ✓ | ✓ | ✓ |  |  |  | ✓ |
| P337 | 6 | ✓ | ✓ |  |  | ✓ | ✓ | ✓ |  |  | ✓ |  |  |
| P338 | 6 |  | ✓ | ✓ |  | ✓ |  |  | ✓ | ✓ | ✓ |  |  |
| P339 | 6 |  | ✓ | ✓ | ✓ |  | ✓ |  | ✓ |  | ✓ |  |  |
| P340 | 6 |  | ✓ |  | ✓ | ✓ |  |  |  | ✓ | ✓ |  | ✓ |
| P341 | 6 |  |  | ✓ | ✓ |  |  | ✓ | ✓ |  |  | ✓ | ✓ |
| P342 | 6 |  |  |  | ✓ | ✓ | ✓ | ✓ | ✓ |  | ✓ |  |  |
| P343 | 6 | ✓ | ✓ | ✓ | ✓ |  |  |  |  | ✓ | ✓ |  |  |
| P344 | 6 | ✓ |  | ✓ |  |  | ✓ | ✓ |  |  | ✓ | ✓ |  |
| P345 | 6 | ✓ | ✓ |  | ✓ |  |  |  | ✓ |  | ✓ |  | ✓ |
| P346 | 6 |  |  | ✓ |  |  | ✓ | ✓ | ✓ | ✓ |  | ✓ |  |
| P347 | 6 | ✓ |  | ✓ | ✓ | ✓ |  |  | ✓ |  |  | ✓ |  |
| P348 | 6 | ✓ | ✓ |  |  | ✓ |  |  | ✓ | ✓ | ✓ |  |  |
| P349 | 6 | ✓ |  | ✓ | ✓ | ✓ |  | ✓ | ✓ |  |  |  |  |
| P350 | 6 | ✓ |  | ✓ |  | ✓ |  | ✓ |  |  | ✓ |  | ✓ |
| P351 | 6 |  | ✓ | ✓ |  |  |  | ✓ |  | ✓ | ✓ |  | ✓ |
| P352 | 6 | ✓ | ✓ | ✓ |  |  |  | ✓ |  |  | ✓ | ✓ |  |
| P353 | 6 |  | ✓ | ✓ |  |  |  |  |  | ✓ | ✓ | ✓ | ✓ |
| P354 | 6 |  | ✓ | ✓ |  |  |  | ✓ | ✓ | ✓ |  | ✓ |  |
| P355 | 6 | ✓ | ✓ |  |  | ✓ | ✓ | ✓ | ✓ |  |  |  |  |
| P356 | 6 |  |  | ✓ | ✓ | ✓ | ✓ |  | ✓ |  |  | ✓ |  |
| P357 | 6 |  |  |  | ✓ | ✓ |  | ✓ | ✓ |  | ✓ |  | ✓ |
| P358 | 6 |  |  |  | ✓ | ✓ | ✓ |  |  | ✓ |  | ✓ | ✓ |
| P359 | 6 | ✓ | ✓ |  |  | ✓ | ✓ |  |  |  | ✓ | ✓ |  |
| P360 | 6 | ✓ |  | ✓ |  | ✓ |  |  | ✓ | ✓ |  |  | ✓ |
| P361 | 6 | ✓ |  | ✓ |  | ✓ | ✓ |  |  | ✓ |  | ✓ |  |
| P362 | 6 |  |  |  |  |  | ✓ |  | ✓ | ✓ | ✓ | ✓ | ✓ |
| P363 | 6 | ✓ | ✓ | ✓ |  | ✓ |  |  | ✓ |  | ✓ |  |  |
| P364 | 6 | ✓ |  |  |  | ✓ |  | ✓ | ✓ |  | ✓ |  | ✓ |
| P365 | 6 | ✓ | ✓ |  | ✓ | ✓ | ✓ |  |  |  |  |  | ✓ |
| P366 | 6 | ✓ |  | ✓ |  |  | ✓ |  | ✓ |  | ✓ |  | ✓ |
| P367 | 6 | ✓ |  | ✓ | ✓ | ✓ |  |  | ✓ |  | ✓ |  |  |
| P368 | 6 |  | ✓ | ✓ |  | ✓ | ✓ |  |  | ✓ | ✓ |  |  |
| P369 | 6 | ✓ | ✓ |  | ✓ |  | ✓ |  |  | ✓ |  | ✓ |  |
| P370 | 6 |  | ✓ |  | ✓ | ✓ |  | ✓ | ✓ |  |  | ✓ |  |
| P371 | 6 |  |  | ✓ | ✓ |  |  | ✓ | ✓ |  | ✓ |  | ✓ |
| P372 | 6 |  |  | ✓ | ✓ | ✓ |  |  |  | ✓ | ✓ |  | ✓ |
| P373 | 6 | ✓ |  |  |  |  | ✓ |  |  | ✓ | ✓ | ✓ | ✓ |
| P374 | 6 |  | ✓ |  | ✓ | ✓ | ✓ |  |  | ✓ | ✓ |  |  |
| P375 | 6 | ✓ | ✓ | ✓ |  | ✓ |  | ✓ |  |  |  |  | ✓ |
| P376 | 6 | ✓ | ✓ |  |  | ✓ |  | ✓ | ✓ |  |  |  | ✓ |
| P377 | 6 |  |  | ✓ | ✓ |  | ✓ | ✓ | ✓ |  |  | ✓ |  |
| P378 | 6 | ✓ | ✓ |  |  |  |  |  | ✓ |  | ✓ | ✓ | ✓ |
| P379 | 6 |  |  |  |  | ✓ |  | ✓ | ✓ | ✓ | ✓ |  | ✓ |
| P380 | 6 |  |  |  |  |  | ✓ | ✓ | ✓ | ✓ | ✓ |  | ✓ |
| P381 | 6 | ✓ | ✓ | ✓ |  | ✓ | ✓ |  |  |  |  |  | ✓ |
| P382 | 6 |  | ✓ | ✓ | ✓ |  |  |  |  | ✓ | ✓ |  | ✓ |
| P383 | 6 | ✓ | ✓ | ✓ | ✓ |  | ✓ |  |  |  |  |  | ✓ |
| P384 | 6 | ✓ | ✓ | ✓ |  |  |  | ✓ |  | ✓ | ✓ |  |  |
| P385 | 6 |  |  | ✓ | ✓ |  | ✓ | ✓ |  | ✓ |  | ✓ |  |
| P386 | 6 | ✓ |  |  |  | ✓ | ✓ | ✓ | ✓ |  |  | ✓ |  |
| P387 | 6 | ✓ |  | ✓ |  |  | ✓ | ✓ |  | ✓ |  |  | ✓ |
| P388 | 6 | ✓ |  | ✓ |  | ✓ | ✓ |  |  | ✓ |  |  | ✓ |
| P389 | 6 | ✓ | ✓ |  | ✓ | ✓ | ✓ |  |  |  |  | ✓ |  |
| P390 | 6 |  | ✓ | ✓ |  | ✓ |  | ✓ | ✓ |  |  |  | ✓ |
| P391 | 6 | ✓ |  | ✓ |  | ✓ |  | ✓ |  |  |  | ✓ | ✓ |
| P392 | 6 | ✓ |  |  |  | ✓ | ✓ |  |  | ✓ | ✓ |  | ✓ |
| P393 | 6 | ✓ |  | ✓ |  | ✓ |  | ✓ |  | ✓ | ✓ |  |  |
| P394 | 6 |  | ✓ |  |  | ✓ | ✓ |  | ✓ | ✓ |  |  | ✓ |
| P395 | 6 | ✓ | ✓ |  | ✓ |  |  | ✓ | ✓ | ✓ |  |  |  |
| P396 | 6 | ✓ | ✓ |  | ✓ |  | ✓ | ✓ |  |  |  |  | ✓ |
| P397 | 6 | ✓ | ✓ |  |  | ✓ |  | ✓ | ✓ |  |  | ✓ |  |
| P398 | 6 | ✓ | ✓ | ✓ |  |  | ✓ |  |  |  |  | ✓ | ✓ |
| P399 | 6 |  | ✓ | ✓ | ✓ | ✓ |  |  |  |  |  | ✓ | ✓ |
| P400 | 6 |  | ✓ | ✓ | ✓ | ✓ |  |  | ✓ |  |  |  | ✓ |
| P401 | 6 |  | ✓ | ✓ | ✓ | ✓ |  |  |  | ✓ |  |  | ✓ |
| P402 | 6 |  | ✓ |  |  |  |  | ✓ | ✓ | ✓ |  | ✓ | ✓ |
| P403 | 6 | ✓ | ✓ | ✓ |  |  |  | ✓ | ✓ |  |  | ✓ |  |
| P404 | 6 |  | ✓ |  |  | ✓ | ✓ | ✓ |  | ✓ | ✓ |  |  |
| P405 | 6 | ✓ | ✓ |  | ✓ | ✓ | ✓ | ✓ |  |  |  |  |  |
| P406 | 6 | ✓ | ✓ |  |  | ✓ |  | ✓ |  |  |  | ✓ | ✓ |
| P407 | 6 |  | ✓ |  | ✓ |  |  | ✓ | ✓ |  | ✓ |  | ✓ |
| P408 | 6 | ✓ | ✓ | ✓ |  |  | ✓ | ✓ |  | ✓ |  |  |  |
| P409 | 6 | ✓ | ✓ | ✓ |  |  |  | ✓ |  |  |  | ✓ | ✓ |
| P410 | 6 | ✓ | ✓ |  |  |  | ✓ | ✓ | ✓ |  |  | ✓ |  |
| P411 | 6 | ✓ |  |  | ✓ | ✓ |  |  | ✓ | ✓ | ✓ |  |  |
| P412 | 6 | ✓ |  | ✓ | ✓ |  | ✓ | ✓ |  |  |  |  | ✓ |
| P413 | 6 |  | ✓ |  | ✓ | ✓ |  | ✓ |  | ✓ |  |  | ✓ |
| P414 | 6 | ✓ |  | ✓ |  |  |  | ✓ |  | ✓ |  | ✓ | ✓ |
| P415 | 6 |  |  | ✓ | ✓ | ✓ | ✓ |  |  | ✓ |  |  | ✓ |
| P416 | 6 | ✓ | ✓ |  | ✓ |  |  | ✓ |  | ✓ |  |  | ✓ |
| P417 | 6 | ✓ |  | ✓ | ✓ | ✓ | ✓ |  | ✓ |  |  |  |  |
| P418 | 6 |  | ✓ |  |  | ✓ |  | ✓ |  | ✓ | ✓ |  | ✓ |
| P419 | 6 |  |  | ✓ |  | ✓ | ✓ | ✓ | ✓ | ✓ |  |  |  |
| P420 | 6 | ✓ |  | ✓ |  |  | ✓ | ✓ |  |  | ✓ |  | ✓ |
| P421 | 6 |  |  | ✓ | ✓ | ✓ |  |  | ✓ |  | ✓ |  | ✓ |
| P422 | 6 | ✓ |  | ✓ | ✓ |  | ✓ |  |  | ✓ |  |  | ✓ |
| P423 | 6 |  |  | ✓ |  | ✓ | ✓ | ✓ |  |  | ✓ | ✓ |  |
| P424 | 6 | ✓ | ✓ |  | ✓ |  | ✓ |  | ✓ |  |  | ✓ |  |
| P425 | 6 |  | ✓ |  | ✓ | ✓ |  |  | ✓ |  |  | ✓ | ✓ |
| P426 | 6 |  |  | ✓ |  |  | ✓ | ✓ |  | ✓ | ✓ |  | ✓ |
| P427 | 6 | ✓ | ✓ |  |  | ✓ | ✓ |  |  | ✓ |  | ✓ |  |
| P428 | 6 |  | ✓ |  |  | ✓ | ✓ | ✓ |  |  | ✓ | ✓ |  |
| P429 | 6 |  | ✓ |  | ✓ | ✓ |  |  | ✓ | ✓ | ✓ |  |  |
| P430 | 6 |  |  | ✓ | ✓ |  | ✓ |  |  | ✓ | ✓ |  | ✓ |
| P431 | 6 | ✓ | ✓ |  |  | ✓ |  | ✓ |  | ✓ | ✓ |  |  |
| P432 | 6 | ✓ |  |  | ✓ |  | ✓ |  |  |  | ✓ | ✓ | ✓ |
| P433 | 6 | ✓ | ✓ |  |  | ✓ | ✓ |  |  |  |  | ✓ | ✓ |
| P434 | 6 |  |  |  |  | ✓ | ✓ | ✓ | ✓ |  | ✓ | ✓ |  |
| P435 | 6 | ✓ | ✓ | ✓ | ✓ | ✓ |  |  |  |  | ✓ |  |  |
| P436 | 6 | ✓ |  | ✓ |  |  |  | ✓ | ✓ |  | ✓ |  | ✓ |
| P437 | 6 |  |  | ✓ | ✓ |  |  |  |  | ✓ | ✓ | ✓ | ✓ |
| P438 | 6 | ✓ |  | ✓ |  | ✓ | ✓ |  |  |  | ✓ |  | ✓ |
| P439 | 6 |  | ✓ | ✓ |  |  | ✓ |  | ✓ |  | ✓ | ✓ |  |
| P440 | 6 | ✓ |  | ✓ | ✓ |  | ✓ |  | ✓ |  | ✓ |  |  |
| P441 | 6 |  |  | ✓ | ✓ |  | ✓ |  | ✓ |  | ✓ |  | ✓ |
| P442 | 6 | ✓ |  |  | ✓ | ✓ | ✓ | ✓ |  |  | ✓ |  |  |
| P443 | 6 |  |  |  | ✓ | ✓ | ✓ |  | ✓ | ✓ |  | ✓ |  |
| P444 | 6 | ✓ | ✓ |  | ✓ |  |  |  |  | ✓ | ✓ | ✓ |  |
| P445 | 6 | ✓ |  |  |  |  | ✓ |  | ✓ |  | ✓ | ✓ | ✓ |
| P446 | 6 |  |  | ✓ | ✓ | ✓ |  | ✓ |  |  | ✓ |  | ✓ |
| P447 | 6 |  | ✓ | ✓ |  |  | ✓ | ✓ |  | ✓ |  |  | ✓ |
| P448 | 6 |  | ✓ |  | ✓ | ✓ | ✓ | ✓ | ✓ |  |  |  |  |
| P449 | 6 | ✓ | ✓ | ✓ |  |  |  |  |  | ✓ |  | ✓ | ✓ |
| P450 | 6 |  | ✓ |  |  |  | ✓ | ✓ | ✓ | ✓ |  |  | ✓ |
| P451 | 6 |  |  | ✓ |  | ✓ | ✓ | ✓ |  | ✓ |  | ✓ |  |
| P452 | 6 | ✓ | ✓ |  |  |  |  | ✓ | ✓ | ✓ | ✓ |  |  |
| P453 | 6 |  |  | ✓ | ✓ |  | ✓ |  |  |  | ✓ | ✓ | ✓ |
| P454 | 6 |  | ✓ |  |  | ✓ |  | ✓ | ✓ | ✓ |  | ✓ |  |
| P455 | 6 |  |  | ✓ |  |  | ✓ |  |  | ✓ | ✓ | ✓ | ✓ |
| P456 | 6 |  |  |  | ✓ | ✓ | ✓ |  |  | ✓ | ✓ | ✓ |  |
| P457 | 6 |  | ✓ | ✓ | ✓ |  |  | ✓ | ✓ |  | ✓ |  |  |
| P458 | 6 | ✓ |  |  | ✓ | ✓ | ✓ |  | ✓ |  |  | ✓ |  |
| P459 | 6 |  |  | ✓ |  | ✓ |  | ✓ | ✓ | ✓ |  |  | ✓ |
| P460 | 6 | ✓ |  |  |  |  |  | ✓ | ✓ | ✓ |  | ✓ | ✓ |
| P461 | 6 | ✓ | ✓ | ✓ | ✓ |  |  |  | ✓ |  | ✓ |  |  |
| P462 | 6 |  |  |  |  |  | ✓ | ✓ | ✓ | ✓ |  | ✓ | ✓ |
| P463 | 6 | ✓ | ✓ |  |  | ✓ |  | ✓ |  |  | ✓ |  | ✓ |
| P464 | 6 | ✓ | ✓ | ✓ |  |  | ✓ | ✓ |  |  | ✓ |  |  |
| P465 | 6 |  | ✓ | ✓ |  | ✓ |  | ✓ | ✓ | ✓ |  |  |  |
| P466 | 6 | ✓ | ✓ |  |  |  | ✓ | ✓ | ✓ |  | ✓ |  |  |
| P467 | 6 |  | ✓ | ✓ |  |  | ✓ | ✓ | ✓ | ✓ |  |  |  |
| P468 | 6 | ✓ | ✓ | ✓ |  |  |  |  |  |  | ✓ | ✓ | ✓ |
| P469 | 6 |  | ✓ | ✓ |  | ✓ | ✓ | ✓ |  |  |  |  | ✓ |
| P470 | 6 |  | ✓ |  | ✓ |  | ✓ | ✓ |  |  |  | ✓ | ✓ |
| P471 | 6 | ✓ |  |  | ✓ |  |  | ✓ | ✓ | ✓ | ✓ |  |  |
| P472 | 6 | ✓ |  | ✓ |  |  | ✓ | ✓ | ✓ | ✓ |  |  |  |
| P473 | 6 |  | ✓ |  | ✓ | ✓ | ✓ |  | ✓ |  |  |  | ✓ |
| P474 | 6 |  | ✓ |  |  |  | ✓ |  | ✓ | ✓ |  | ✓ | ✓ |
| P475 | 6 |  |  | ✓ |  | ✓ | ✓ |  |  | ✓ | ✓ |  | ✓ |
| P476 | 6 |  |  | ✓ | ✓ |  |  | ✓ |  |  | ✓ | ✓ | ✓ |
| P477 | 6 | ✓ |  |  |  | ✓ |  | ✓ | ✓ |  | ✓ | ✓ |  |
| P478 | 6 |  | ✓ |  |  |  |  | ✓ | ✓ | ✓ | ✓ |  | ✓ |
| P479 | 6 |  | ✓ |  |  |  |  | ✓ | ✓ |  | ✓ | ✓ | ✓ |
| P480 | 6 | ✓ |  |  |  | ✓ |  | ✓ | ✓ | ✓ |  | ✓ |  |
| P481 | 6 |  | ✓ |  |  | ✓ | ✓ |  | ✓ |  | ✓ | ✓ |  |
| P482 | 6 | ✓ |  | ✓ | ✓ |  |  | ✓ | ✓ |  |  |  | ✓ |
| P483 | 6 | ✓ |  |  |  | ✓ |  | ✓ | ✓ | ✓ |  |  | ✓ |
| P484 | 6 | ✓ |  |  | ✓ |  |  | ✓ |  | ✓ | ✓ | ✓ |  |
| P485 | 6 |  |  |  | ✓ | ✓ |  | ✓ |  | ✓ |  | ✓ | ✓ |
| P486 | 6 | ✓ |  |  | ✓ |  | ✓ | ✓ |  |  |  | ✓ | ✓ |
| P487 | 6 |  | ✓ | ✓ |  | ✓ |  |  | ✓ | ✓ |  | ✓ |  |
| P488 | 7 | ✓ | ✓ | ✓ | ✓ |  |  | ✓ | ✓ |  |  |  | ✓ |
| P489 | 7 |  | ✓ | ✓ | ✓ |  |  | ✓ | ✓ |  | ✓ |  | ✓ |
| P490 | 7 |  | ✓ | ✓ | ✓ | ✓ | ✓ |  |  | ✓ |  | ✓ |  |
| P491 | 7 | ✓ | ✓ |  |  | ✓ | ✓ | ✓ | ✓ |  |  |  | ✓ |
| P492 | 7 | ✓ | ✓ | ✓ |  |  |  |  | ✓ |  | ✓ | ✓ | ✓ |
| P493 | 7 | ✓ |  | ✓ | ✓ | ✓ | ✓ |  |  |  | ✓ | ✓ |  |
| P494 | 7 |  |  |  | ✓ | ✓ |  | ✓ | ✓ |  | ✓ | ✓ | ✓ |
| P495 | 7 | ✓ | ✓ |  | ✓ |  |  | ✓ | ✓ |  | ✓ |  | ✓ |
| P496 | 7 |  | ✓ | ✓ |  |  | ✓ | ✓ | ✓ |  |  | ✓ | ✓ |
| P497 | 7 | ✓ | ✓ |  |  |  |  |  | ✓ | ✓ | ✓ | ✓ | ✓ |
| P498 | 7 | ✓ |  |  |  |  | ✓ | ✓ | ✓ | ✓ |  | ✓ | ✓ |
| P499 | 7 | ✓ | ✓ |  | ✓ | ✓ |  |  | ✓ | ✓ | ✓ |  |  |
| P500 | 7 | ✓ | ✓ | ✓ | ✓ |  |  |  |  | ✓ | ✓ |  | ✓ |
| P501 | 7 |  | ✓ | ✓ |  | ✓ | ✓ | ✓ |  | ✓ |  | ✓ |  |
| P502 | 7 | ✓ | ✓ |  |  |  | ✓ | ✓ | ✓ |  | ✓ | ✓ |  |
| P503 | 7 | ✓ |  | ✓ | ✓ |  | ✓ | ✓ | ✓ |  | ✓ |  |  |
| P504 | 7 | ✓ | ✓ |  | ✓ |  |  | ✓ | ✓ |  | ✓ | ✓ |  |
| P505 | 7 |  |  |  | ✓ | ✓ | ✓ | ✓ |  | ✓ |  | ✓ | ✓ |
| P506 | 7 | ✓ |  |  | ✓ | ✓ |  |  |  | ✓ | ✓ | ✓ | ✓ |
| P507 | 7 |  | ✓ |  | ✓ |  | ✓ | ✓ | ✓ |  | ✓ | ✓ |  |
| P508 | 7 |  |  |  |  | ✓ |  | ✓ | ✓ | ✓ | ✓ | ✓ | ✓ |
| P509 | 7 | ✓ |  | ✓ | ✓ |  |  | ✓ | ✓ | ✓ |  | ✓ |  |
| P510 | 7 |  |  |  | ✓ |  | ✓ | ✓ | ✓ | ✓ |  | ✓ | ✓ |
| P511 | 7 | ✓ | ✓ |  |  | ✓ | ✓ | ✓ |  |  | ✓ |  | ✓ |
| P512 | 7 |  | ✓ | ✓ |  | ✓ | ✓ | ✓ |  | ✓ | ✓ |  |  |
| P513 | 7 |  | ✓ | ✓ |  | ✓ |  |  | ✓ | ✓ | ✓ |  | ✓ |
| P514 | 7 | ✓ |  |  | ✓ | ✓ | ✓ |  | ✓ | ✓ |  |  | ✓ |
| P515 | 7 | ✓ |  |  | ✓ | ✓ |  |  | ✓ | ✓ | ✓ | ✓ |  |
| P516 | 7 |  |  | ✓ | ✓ |  |  | ✓ |  | ✓ | ✓ | ✓ | ✓ |
| P517 | 7 | ✓ |  | ✓ |  | ✓ |  | ✓ | ✓ |  | ✓ | ✓ |  |
| P518 | 7 | ✓ | ✓ |  | ✓ | ✓ |  |  |  | ✓ |  | ✓ | ✓ |
| P519 | 7 |  | ✓ | ✓ | ✓ | ✓ | ✓ |  | ✓ | ✓ |  |  |  |
| P520 | 7 |  | ✓ | ✓ | ✓ |  | ✓ |  | ✓ | ✓ |  | ✓ |  |
| P521 | 7 | ✓ | ✓ | ✓ |  | ✓ | ✓ | ✓ | ✓ |  |  |  |  |
| P522 | 7 | ✓ |  |  |  |  | ✓ | ✓ | ✓ | ✓ | ✓ |  | ✓ |
| P523 | 7 |  | ✓ |  | ✓ | ✓ |  | ✓ |  | ✓ |  | ✓ | ✓ |
| P524 | 7 | ✓ |  | ✓ |  | ✓ |  |  |  | ✓ | ✓ | ✓ | ✓ |
| P525 | 7 | ✓ |  | ✓ | ✓ | ✓ |  |  | ✓ | ✓ |  | ✓ |  |
| P526 | 7 | ✓ |  | ✓ | ✓ | ✓ |  | ✓ | ✓ |  |  | ✓ |  |
| P527 | 7 |  | ✓ |  |  | ✓ | ✓ | ✓ | ✓ | ✓ |  | ✓ |  |
| P528 | 7 | ✓ |  | ✓ |  | ✓ |  | ✓ | ✓ |  | ✓ |  | ✓ |
| P529 | 7 | ✓ | ✓ |  | ✓ | ✓ | ✓ | ✓ |  |  | ✓ |  |  |
| P530 | 7 | ✓ | ✓ |  |  | ✓ |  | ✓ | ✓ | ✓ |  | ✓ |  |
| P531 | 7 |  | ✓ |  | ✓ | ✓ |  | ✓ |  |  | ✓ | ✓ | ✓ |
| P532 | 7 |  |  | ✓ | ✓ | ✓ |  | ✓ | ✓ |  |  | ✓ | ✓ |
| P533 | 7 | ✓ |  | ✓ |  | ✓ | ✓ |  | ✓ |  | ✓ |  | ✓ |
| P534 | 7 | ✓ | ✓ | ✓ | ✓ |  | ✓ | ✓ |  |  |  |  | ✓ |
| P535 | 7 |  | ✓ | ✓ |  | ✓ | ✓ | ✓ |  | ✓ |  |  | ✓ |
| P536 | 7 |  | ✓ | ✓ | ✓ |  |  | ✓ | ✓ | ✓ |  | ✓ |  |
| P537 | 7 |  | ✓ |  |  |  | ✓ | ✓ | ✓ | ✓ | ✓ | ✓ |  |
| P538 | 7 |  | ✓ | ✓ |  | ✓ |  | ✓ | ✓ | ✓ | ✓ |  |  |
| P539 | 7 |  |  | ✓ | ✓ |  |  | ✓ | ✓ | ✓ | ✓ |  | ✓ |
| P540 | 7 | ✓ |  |  | ✓ | ✓ | ✓ |  | ✓ | ✓ |  | ✓ |  |
| P541 | 7 |  | ✓ | ✓ | ✓ |  | ✓ | ✓ | ✓ | ✓ |  |  |  |
| P542 | 7 | ✓ | ✓ |  |  | ✓ |  | ✓ | ✓ |  |  | ✓ | ✓ |
| P543 | 7 |  | ✓ |  |  |  | ✓ |  | ✓ | ✓ | ✓ | ✓ | ✓ |
| P544 | 7 |  | ✓ |  |  | ✓ | ✓ |  | ✓ | ✓ |  | ✓ | ✓ |
| P545 | 7 | ✓ |  |  | ✓ |  | ✓ | ✓ |  | ✓ | ✓ |  | ✓ |
| P546 | 7 |  |  |  | ✓ | ✓ | ✓ | ✓ | ✓ | ✓ |  |  | ✓ |
| P547 | 7 |  | ✓ |  |  | ✓ |  | ✓ | ✓ | ✓ | ✓ | ✓ |  |
| P548 | 7 | ✓ | ✓ |  | ✓ |  | ✓ | ✓ | ✓ |  | ✓ |  |  |
| P549 | 7 | ✓ | ✓ |  |  |  | ✓ | ✓ |  | ✓ | ✓ | ✓ |  |
| P550 | 7 |  |  | ✓ | ✓ | ✓ | ✓ | ✓ | ✓ | ✓ |  |  |  |
| P551 | 7 |  | ✓ | ✓ |  |  | ✓ |  | ✓ | ✓ | ✓ | ✓ |  |
| P552 | 7 |  | ✓ | ✓ | ✓ |  | ✓ |  |  | ✓ |  | ✓ | ✓ |
| P553 | 7 | ✓ |  | ✓ |  | ✓ | ✓ | ✓ |  |  | ✓ | ✓ |  |
| P554 | 7 | ✓ | ✓ |  |  | ✓ |  | ✓ |  | ✓ |  | ✓ | ✓ |
| P555 | 7 | ✓ |  |  | ✓ | ✓ | ✓ |  | ✓ | ✓ | ✓ |  |  |
| P556 | 7 | ✓ | ✓ | ✓ |  |  |  | ✓ | ✓ |  |  | ✓ | ✓ |
| P557 | 7 |  |  |  | ✓ | ✓ | ✓ |  | ✓ | ✓ | ✓ |  | ✓ |
| P558 | 7 |  | ✓ |  |  | ✓ | ✓ | ✓ | ✓ |  | ✓ | ✓ |  |
| P559 | 7 | ✓ | ✓ | ✓ | ✓ |  |  |  |  | ✓ |  | ✓ | ✓ |
| P560 | 7 | ✓ | ✓ |  |  | ✓ |  | ✓ | ✓ | ✓ | ✓ |  |  |
| P561 | 7 |  | ✓ |  | ✓ | ✓ | ✓ | ✓ |  |  | ✓ |  | ✓ |
| P562 | 7 |  | ✓ | ✓ |  | ✓ |  | ✓ | ✓ |  | ✓ |  | ✓ |
| P563 | 7 |  | ✓ |  |  | ✓ | ✓ |  | ✓ |  | ✓ | ✓ | ✓ |
| P564 | 7 |  |  | ✓ | ✓ | ✓ | ✓ | ✓ |  |  |  | ✓ | ✓ |
| P565 | 7 | ✓ |  |  | ✓ |  | ✓ | ✓ | ✓ | ✓ |  | ✓ |  |
| P566 | 7 | ✓ | ✓ |  | ✓ | ✓ | ✓ |  | ✓ |  |  | ✓ |  |
| P567 | 7 |  |  | ✓ | ✓ |  | ✓ | ✓ | ✓ |  | ✓ | ✓ |  |
| P568 | 7 |  |  | ✓ | ✓ | ✓ | ✓ |  | ✓ |  |  | ✓ | ✓ |
| P569 | 7 | ✓ |  |  |  | ✓ | ✓ |  | ✓ | ✓ |  | ✓ | ✓ |
| P570 | 7 | ✓ | ✓ | ✓ |  |  | ✓ |  | ✓ | ✓ |  |  | ✓ |
| P571 | 7 |  |  |  | ✓ | ✓ | ✓ | ✓ | ✓ |  | ✓ |  | ✓ |
| P572 | 7 | ✓ | ✓ |  |  | ✓ | ✓ |  | ✓ |  | ✓ | ✓ |  |
| P573 | 7 | ✓ |  | ✓ |  | ✓ |  | ✓ |  | ✓ | ✓ | ✓ |  |
| P574 | 7 | ✓ |  |  | ✓ | ✓ |  | ✓ | ✓ | ✓ |  |  | ✓ |
| P575 | 7 | ✓ |  | ✓ | ✓ | ✓ | ✓ |  | ✓ | ✓ |  |  |  |
| P576 | 7 | ✓ | ✓ |  | ✓ |  | ✓ |  | ✓ |  |  | ✓ | ✓ |
| P577 | 7 | ✓ |  |  | ✓ | ✓ | ✓ | ✓ |  |  | ✓ | ✓ |  |
| P578 | 7 | ✓ | ✓ |  | ✓ | ✓ | ✓ |  |  |  |  | ✓ | ✓ |
| P579 | 7 | ✓ | ✓ | ✓ | ✓ |  | ✓ | ✓ |  |  | ✓ |  |  |
| P580 | 7 |  | ✓ | ✓ | ✓ | ✓ | ✓ | ✓ |  |  |  | ✓ |  |
| P581 | 7 | ✓ | ✓ |  | ✓ | ✓ | ✓ | ✓ |  |  |  | ✓ |  |
| P582 | 7 | ✓ | ✓ |  | ✓ |  | ✓ | ✓ | ✓ |  |  |  | ✓ |
| P583 | 7 |  | ✓ | ✓ |  |  |  |  | ✓ | ✓ | ✓ | ✓ | ✓ |
| P584 | 7 | ✓ |  | ✓ |  |  |  | ✓ |  | ✓ | ✓ | ✓ | ✓ |
| P585 | 7 | ✓ | ✓ |  | ✓ |  | ✓ | ✓ |  |  | ✓ |  | ✓ |
| P586 | 7 | ✓ |  |  |  | ✓ | ✓ |  | ✓ | ✓ | ✓ | ✓ |  |
| P587 | 7 | ✓ | ✓ |  |  |  | ✓ | ✓ | ✓ | ✓ |  |  | ✓ |
| P588 | 7 | ✓ |  |  | ✓ |  | ✓ | ✓ |  | ✓ |  | ✓ | ✓ |
| P589 | 7 |  | ✓ | ✓ | ✓ |  | ✓ |  | ✓ | ✓ | ✓ |  |  |
| P590 | 7 | ✓ | ✓ | ✓ |  | ✓ | ✓ |  | ✓ |  |  | ✓ |  |
| P591 | 7 | ✓ | ✓ | ✓ |  | ✓ |  | ✓ |  |  | ✓ |  | ✓ |
| P592 | 7 |  | ✓ | ✓ | ✓ | ✓ |  | ✓ |  | ✓ |  | ✓ |  |
| P593 | 7 |  | ✓ | ✓ |  | ✓ | ✓ |  | ✓ |  |  | ✓ | ✓ |
| P594 | 7 |  | ✓ | ✓ | ✓ | ✓ | ✓ |  |  |  | ✓ | ✓ |  |
| P595 | 7 | ✓ |  | ✓ | ✓ |  | ✓ | ✓ | ✓ |  |  |  | ✓ |
| P596 | 7 | ✓ |  | ✓ |  | ✓ |  | ✓ | ✓ | ✓ | ✓ |  |  |
| P597 | 7 | ✓ |  | ✓ |  | ✓ | ✓ | ✓ | ✓ | ✓ |  |  |  |
| P598 | 7 |  | ✓ | ✓ | ✓ | ✓ |  | ✓ | ✓ |  | ✓ |  |  |
| P599 | 7 |  | ✓ | ✓ |  | ✓ |  | ✓ |  | ✓ | ✓ |  | ✓ |
| P600 | 7 | ✓ |  |  |  | ✓ |  | ✓ | ✓ | ✓ | ✓ | ✓ |  |
| P601 | 7 | ✓ | ✓ | ✓ |  |  | ✓ |  |  |  | ✓ | ✓ | ✓ |
| P602 | 7 | ✓ |  | ✓ | ✓ |  |  | ✓ |  |  | ✓ | ✓ | ✓ |
| P603 | 7 |  |  | ✓ |  | ✓ | ✓ | ✓ |  |  | ✓ | ✓ | ✓ |
| P604 | 7 | ✓ |  | ✓ | ✓ | ✓ |  |  |  |  | ✓ | ✓ | ✓ |
| P605 | 7 |  | ✓ | ✓ |  |  |  | ✓ | ✓ | ✓ |  | ✓ | ✓ |
| P606 | 7 | ✓ |  | ✓ | ✓ |  | ✓ | ✓ |  | ✓ | ✓ |  |  |
| P607 | 7 | ✓ | ✓ |  |  | ✓ |  | ✓ |  |  | ✓ | ✓ | ✓ |
| P608 | 7 | ✓ |  | ✓ | ✓ |  | ✓ | ✓ |  |  |  | ✓ | ✓ |
| P609 | 7 |  | ✓ |  | ✓ | ✓ | ✓ |  |  | ✓ | ✓ | ✓ |  |
| P610 | 7 |  | ✓ |  | ✓ |  | ✓ |  | ✓ |  | ✓ | ✓ | ✓ |
| P611 | 7 | ✓ | ✓ | ✓ | ✓ |  |  | ✓ | ✓ |  |  | ✓ |  |
| P612 | 7 | ✓ | ✓ |  |  |  | ✓ | ✓ | ✓ |  |  | ✓ | ✓ |
| P613 | 7 |  |  | ✓ |  | ✓ |  | ✓ |  | ✓ | ✓ | ✓ | ✓ |
| P614 | 7 |  | ✓ |  | ✓ | ✓ |  |  | ✓ | ✓ | ✓ | ✓ |  |
| P615 | 7 | ✓ | ✓ | ✓ | ✓ |  | ✓ |  |  | ✓ |  |  | ✓ |
| P616 | 7 | ✓ | ✓ | ✓ |  | ✓ |  |  | ✓ | ✓ |  | ✓ |  |
| P617 | 7 | ✓ |  |  | ✓ | ✓ | ✓ |  |  | ✓ | ✓ |  | ✓ |
| P618 | 7 | ✓ |  | ✓ | ✓ | ✓ |  | ✓ |  |  |  | ✓ | ✓ |
| P619 | 7 | ✓ | ✓ |  |  | ✓ | ✓ |  | ✓ | ✓ |  |  | ✓ |
| P620 | 7 | ✓ | ✓ | ✓ |  | ✓ |  | ✓ |  | ✓ | ✓ |  |  |
| P621 | 7 | ✓ | ✓ | ✓ |  |  | ✓ | ✓ | ✓ |  |  | ✓ |  |
| P622 | 7 | ✓ | ✓ | ✓ | ✓ |  | ✓ |  | ✓ |  |  |  | ✓ |
| P623 | 7 |  |  | ✓ | ✓ |  | ✓ |  | ✓ | ✓ |  | ✓ | ✓ |
| P624 | 7 | ✓ | ✓ |  | ✓ |  | ✓ |  | ✓ |  | ✓ | ✓ |  |
| P625 | 7 | ✓ |  | ✓ |  |  |  | ✓ | ✓ | ✓ | ✓ |  | ✓ |
| P626 | 7 |  |  | ✓ | ✓ | ✓ | ✓ |  | ✓ | ✓ | ✓ |  |  |
| P627 | 7 | ✓ | ✓ | ✓ |  |  |  | ✓ | ✓ |  | ✓ | ✓ |  |
| P628 | 7 |  |  | ✓ | ✓ | ✓ |  | ✓ | ✓ |  | ✓ | ✓ |  |
| P629 | 7 |  | ✓ |  | ✓ | ✓ | ✓ |  |  |  | ✓ | ✓ | ✓ |
| P630 | 7 |  | ✓ | ✓ | ✓ | ✓ | ✓ | ✓ |  |  | ✓ |  |  |
| P631 | 7 | ✓ |  | ✓ |  |  | ✓ | ✓ | ✓ | ✓ | ✓ |  |  |
| P632 | 7 | ✓ |  |  | ✓ |  | ✓ |  | ✓ | ✓ | ✓ | ✓ |  |
| P633 | 7 | ✓ | ✓ | ✓ |  |  | ✓ | ✓ |  | ✓ | ✓ |  |  |
| P634 | 7 |  |  |  | ✓ |  | ✓ | ✓ | ✓ | ✓ | ✓ |  | ✓ |
| P635 | 7 | ✓ |  |  | ✓ | ✓ | ✓ |  |  | ✓ |  | ✓ | ✓ |
| P636 | 7 | ✓ |  |  | ✓ | ✓ |  | ✓ | ✓ |  |  | ✓ | ✓ |
| P637 | 7 | ✓ | ✓ |  | ✓ | ✓ |  | ✓ |  | ✓ |  |  | ✓ |
| P638 | 7 | ✓ |  |  | ✓ | ✓ | ✓ |  | ✓ |  | ✓ | ✓ |  |
| P639 | 7 |  | ✓ |  | ✓ | ✓ | ✓ | ✓ |  | ✓ | ✓ |  |  |
| P640 | 7 |  |  | ✓ | ✓ |  | ✓ | ✓ | ✓ |  |  | ✓ | ✓ |
| P641 | 7 | ✓ |  | ✓ |  | ✓ | ✓ |  | ✓ |  |  | ✓ | ✓ |
| P642 | 7 | ✓ | ✓ |  |  |  | ✓ |  | ✓ | ✓ | ✓ |  | ✓ |
| P643 | 7 | ✓ | ✓ | ✓ |  |  | ✓ |  | ✓ |  | ✓ |  | ✓ |
| P644 | 7 | ✓ | ✓ | ✓ | ✓ | ✓ |  | ✓ |  |  |  |  | ✓ |
| P645 | 7 |  | ✓ |  | ✓ |  | ✓ | ✓ | ✓ | ✓ |  | ✓ |  |
| P646 | 7 | ✓ |  | ✓ |  |  | ✓ | ✓ |  | ✓ | ✓ | ✓ |  |
| P647 | 8 |  | ✓ | ✓ | ✓ |  |  |  | ✓ | ✓ | ✓ | ✓ | ✓ |
| P648 | 8 |  | ✓ | ✓ |  | ✓ |  | ✓ | ✓ | ✓ | ✓ |  | ✓ |
| P649 | 8 |  |  | ✓ | ✓ | ✓ | ✓ |  | ✓ | ✓ | ✓ | ✓ |  |
| P650 | 8 | ✓ | ✓ |  | ✓ | ✓ | ✓ |  | ✓ |  | ✓ |  | ✓ |
| P651 | 8 | ✓ | ✓ | ✓ | ✓ | ✓ |  |  | ✓ |  |  | ✓ | ✓ |
| P652 | 8 |  | ✓ | ✓ |  |  |  | ✓ | ✓ | ✓ | ✓ | ✓ | ✓ |
| P653 | 8 | ✓ | ✓ | ✓ |  |  | ✓ | ✓ |  |  | ✓ | ✓ | ✓ |
| P654 | 8 | ✓ |  | ✓ |  | ✓ | ✓ |  | ✓ | ✓ | ✓ |  | ✓ |
| P655 | 8 | ✓ | ✓ | ✓ |  | ✓ |  | ✓ | ✓ |  | ✓ | ✓ |  |
| P656 | 8 | ✓ | ✓ |  |  | ✓ |  | ✓ |  | ✓ | ✓ | ✓ | ✓ |
| P657 | 8 | ✓ | ✓ | ✓ | ✓ | ✓ |  | ✓ | ✓ | ✓ |  |  |  |
| P658 | 8 | ✓ | ✓ |  |  | ✓ | ✓ |  |  | ✓ | ✓ | ✓ | ✓ |
| P659 | 8 |  | ✓ |  | ✓ | ✓ | ✓ | ✓ | ✓ |  |  | ✓ | ✓ |
| P660 | 8 | ✓ | ✓ |  | ✓ |  |  | ✓ | ✓ | ✓ | ✓ |  | ✓ |
| P661 | 8 |  | ✓ |  | ✓ | ✓ | ✓ | ✓ | ✓ | ✓ |  |  | ✓ |
| P662 | 8 |  |  | ✓ | ✓ | ✓ | ✓ |  | ✓ |  | ✓ | ✓ | ✓ |
| P663 | 8 |  | ✓ | ✓ |  |  | ✓ | ✓ |  | ✓ | ✓ | ✓ | ✓ |
| P664 | 8 | ✓ | ✓ |  | ✓ |  |  | ✓ | ✓ | ✓ | ✓ | ✓ |  |
| P665 | 8 | ✓ | ✓ | ✓ | ✓ | ✓ | ✓ |  |  | ✓ |  |  | ✓ |
| P666 | 8 |  | ✓ |  | ✓ |  | ✓ | ✓ | ✓ | ✓ | ✓ |  | ✓ |
| P667 | 8 |  |  | ✓ |  | ✓ | ✓ | ✓ | ✓ | ✓ | ✓ | ✓ |  |
| P668 | 8 | ✓ | ✓ | ✓ | ✓ |  |  |  | ✓ | ✓ | ✓ | ✓ |  |
| P669 | 8 | ✓ | ✓ | ✓ | ✓ |  | ✓ |  | ✓ |  | ✓ |  | ✓ |
| P670 | 8 | ✓ | ✓ |  |  |  | ✓ | ✓ | ✓ | ✓ | ✓ | ✓ |  |
| P671 | 8 | ✓ | ✓ |  | ✓ |  | ✓ |  | ✓ |  | ✓ | ✓ | ✓ |
| P672 | 8 |  | ✓ |  | ✓ | ✓ |  |  | ✓ | ✓ | ✓ | ✓ | ✓ |
| P673 | 8 |  | ✓ | ✓ | ✓ |  | ✓ | ✓ |  | ✓ | ✓ |  | ✓ |
| P674 | 8 | ✓ | ✓ |  | ✓ | ✓ | ✓ | ✓ |  |  | ✓ | ✓ |  |
| P675 | 8 |  |  | ✓ |  | ✓ | ✓ | ✓ | ✓ | ✓ | ✓ |  | ✓ |
| P676 | 8 |  | ✓ | ✓ | ✓ |  |  | ✓ | ✓ |  | ✓ | ✓ | ✓ |
| P677 | 8 | ✓ | ✓ | ✓ |  |  |  | ✓ | ✓ | ✓ | ✓ |  | ✓ |
| P678 | 8 |  | ✓ | ✓ |  | ✓ | ✓ |  | ✓ | ✓ |  | ✓ | ✓ |
| P679 | 8 |  |  |  | ✓ |  | ✓ | ✓ | ✓ | ✓ | ✓ | ✓ | ✓ |
| P680 | 8 | ✓ |  |  | ✓ | ✓ |  |  | ✓ | ✓ | ✓ | ✓ | ✓ |
| P681 | 8 | ✓ |  | ✓ | ✓ |  | ✓ |  | ✓ |  | ✓ | ✓ | ✓ |
| P682 | 8 |  | ✓ | ✓ | ✓ | ✓ |  | ✓ | ✓ | ✓ |  |  | ✓ |
| P683 | 8 | ✓ |  | ✓ | ✓ |  | ✓ | ✓ | ✓ | ✓ |  |  | ✓ |
| P684 | 8 | ✓ |  | ✓ |  |  | ✓ | ✓ | ✓ | ✓ | ✓ |  | ✓ |
| P685 | 8 | ✓ | ✓ | ✓ | ✓ |  | ✓ |  | ✓ | ✓ |  |  | ✓ |
| P686 | 8 |  | ✓ | ✓ | ✓ | ✓ | ✓ | ✓ | ✓ | ✓ |  |  |  |
| P687 | 8 | ✓ | ✓ | ✓ |  | ✓ |  | ✓ |  |  | ✓ | ✓ | ✓ |
| P688 | 8 | ✓ | ✓ | ✓ |  |  | ✓ | ✓ | ✓ | ✓ |  |  | ✓ |
| P689 | 8 | ✓ |  |  | ✓ | ✓ | ✓ | ✓ |  | ✓ | ✓ | ✓ |  |
| P690 | 8 | ✓ | ✓ | ✓ | ✓ | ✓ | ✓ |  |  | ✓ | ✓ |  |  |
| P691 | 8 | ✓ | ✓ |  | ✓ |  | ✓ | ✓ |  | ✓ |  | ✓ | ✓ |
| P692 | 8 | ✓ |  | ✓ |  | ✓ | ✓ |  | ✓ |  | ✓ | ✓ | ✓ |
| P693 | 8 |  | ✓ | ✓ | ✓ | ✓ | ✓ |  |  | ✓ | ✓ |  | ✓ |
| P694 | 8 |  | ✓ | ✓ |  | ✓ |  | ✓ | ✓ | ✓ | ✓ | ✓ |  |
| P695 | 8 | ✓ | ✓ |  |  |  | ✓ |  | ✓ | ✓ | ✓ | ✓ | ✓ |
| P696 | 8 |  |  | ✓ | ✓ | ✓ | ✓ |  | ✓ | ✓ | ✓ |  | ✓ |
| P697 | 8 | ✓ | ✓ |  | ✓ | ✓ | ✓ |  | ✓ | ✓ | ✓ |  |  |
| P698 | 8 | ✓ |  |  | ✓ | ✓ | ✓ |  |  | ✓ | ✓ | ✓ | ✓ |
| P699 | 8 |  | ✓ | ✓ |  | ✓ | ✓ | ✓ |  | ✓ |  | ✓ | ✓ |
| P700 | 8 |  |  | ✓ | ✓ | ✓ | ✓ | ✓ | ✓ |  | ✓ | ✓ |  |
| P701 | 8 | ✓ | ✓ | ✓ | ✓ | ✓ | ✓ |  | ✓ |  |  | ✓ |  |
| P702 | 8 | ✓ | ✓ | ✓ |  |  | ✓ | ✓ |  | ✓ |  | ✓ | ✓ |
| P703 | 8 | ✓ |  | ✓ | ✓ |  |  | ✓ | ✓ | ✓ | ✓ | ✓ |  |
| P704 | 8 | ✓ | ✓ | ✓ | ✓ | ✓ |  |  | ✓ |  | ✓ | ✓ |  |
| P705 | 8 | ✓ |  |  | ✓ | ✓ | ✓ |  | ✓ | ✓ | ✓ |  | ✓ |
| P706 | 8 | ✓ |  |  |  | ✓ | ✓ | ✓ | ✓ |  | ✓ | ✓ | ✓ |
| P707 | 8 | ✓ | ✓ | ✓ |  | ✓ | ✓ | ✓ | ✓ |  |  |  | ✓ |
| P708 | 8 | ✓ |  | ✓ |  | ✓ |  | ✓ | ✓ | ✓ | ✓ | ✓ |  |
| P709 | 8 | ✓ | ✓ | ✓ |  | ✓ | ✓ | ✓ | ✓ |  |  | ✓ |  |
| P710 | 8 | ✓ |  | ✓ |  | ✓ | ✓ | ✓ | ✓ | ✓ | ✓ |  |  |
| P711 | 8 |  | ✓ | ✓ | ✓ |  | ✓ | ✓ |  |  | ✓ | ✓ | ✓ |
| P712 | 8 | ✓ |  | ✓ | ✓ |  |  | ✓ | ✓ | ✓ | ✓ |  | ✓ |
| P713 | 8 | ✓ |  | ✓ | ✓ | ✓ | ✓ |  | ✓ |  | ✓ | ✓ |  |
| P714 | 8 |  |  | ✓ | ✓ |  | ✓ | ✓ | ✓ | ✓ | ✓ | ✓ |  |
| P715 | 8 | ✓ |  | ✓ | ✓ | ✓ | ✓ | ✓ | ✓ | ✓ |  |  |  |
| P716 | 8 | ✓ |  | ✓ | ✓ | ✓ | ✓ | ✓ |  |  | ✓ |  | ✓ |
| P717 | 8 | ✓ | ✓ |  |  | ✓ | ✓ | ✓ | ✓ | ✓ |  | ✓ |  |
| P718 | 8 |  | ✓ | ✓ | ✓ | ✓ |  | ✓ | ✓ | ✓ |  | ✓ |  |
| P719 | 8 | ✓ | ✓ |  | ✓ | ✓ |  | ✓ |  | ✓ |  | ✓ | ✓ |
| P720 | 8 | ✓ | ✓ | ✓ |  |  | ✓ |  | ✓ |  | ✓ | ✓ | ✓ |
| P721 | 8 |  | ✓ | ✓ | ✓ | ✓ | ✓ |  | ✓ |  | ✓ | ✓ |  |
| P722 | 8 |  |  | ✓ | ✓ |  | ✓ | ✓ | ✓ |  | ✓ | ✓ | ✓ |
| P723 | 8 | ✓ | ✓ | ✓ | ✓ |  |  |  | ✓ | ✓ | ✓ |  | ✓ |
| P724 | 8 | ✓ | ✓ | ✓ |  |  | ✓ | ✓ | ✓ |  | ✓ |  | ✓ |
| P725 | 8 |  |  | ✓ | ✓ | ✓ |  | ✓ | ✓ | ✓ |  | ✓ | ✓ |
| P726 | 8 | ✓ | ✓ | ✓ | ✓ |  | ✓ | ✓ | ✓ | ✓ |  |  |  |
| P727 | 8 | ✓ | ✓ | ✓ | ✓ |  | ✓ |  |  |  | ✓ | ✓ | ✓ |
| P728 | 8 | ✓ |  | ✓ | ✓ | ✓ |  | ✓ |  |  | ✓ | ✓ | ✓ |
| P729 | 8 |  | ✓ | ✓ | ✓ |  | ✓ |  | ✓ |  | ✓ | ✓ | ✓ |
| P730 | 8 | ✓ | ✓ | ✓ |  |  | ✓ |  |  | ✓ | ✓ | ✓ | ✓ |
| P731 | 8 |  | ✓ | ✓ | ✓ | ✓ | ✓ |  |  |  | ✓ | ✓ | ✓ |
| P732 | 8 | ✓ |  |  | ✓ | ✓ | ✓ | ✓ |  |  | ✓ | ✓ | ✓ |
| P733 | 8 |  |  | ✓ | ✓ | ✓ | ✓ | ✓ | ✓ | ✓ |  | ✓ |  |
| P734 | 8 | ✓ | ✓ |  | ✓ | ✓ |  | ✓ |  | ✓ | ✓ | ✓ |  |
| P735 | 8 |  |  | ✓ | ✓ | ✓ | ✓ | ✓ | ✓ |  | ✓ |  | ✓ |
| P736 | 8 | ✓ | ✓ | ✓ |  | ✓ |  |  |  | ✓ | ✓ | ✓ | ✓ |
| P737 | 8 | ✓ |  | ✓ |  | ✓ | ✓ |  |  | ✓ | ✓ | ✓ | ✓ |
| P738 | 8 |  | ✓ | ✓ |  | ✓ | ✓ | ✓ | ✓ | ✓ |  | ✓ |  |
| P739 | 8 | ✓ | ✓ |  | ✓ |  |  | ✓ | ✓ |  | ✓ | ✓ | ✓ |
| P740 | 8 |  |  |  | ✓ | ✓ | ✓ | ✓ | ✓ | ✓ | ✓ |  | ✓ |
| P741 | 8 |  |  | ✓ | ✓ | ✓ |  | ✓ | ✓ |  | ✓ | ✓ | ✓ |
| P742 | 8 | ✓ |  | ✓ | ✓ | ✓ |  |  | ✓ | ✓ | ✓ | ✓ |  |
| P743 | 8 | ✓ | ✓ | ✓ | ✓ | ✓ |  | ✓ |  |  | ✓ |  | ✓ |
| P744 | 8 | ✓ | ✓ |  | ✓ |  | ✓ | ✓ | ✓ |  |  | ✓ | ✓ |
| P745 | 8 | ✓ | ✓ | ✓ | ✓ |  | ✓ |  | ✓ | ✓ |  | ✓ |  |
| P746 | 9 | ✓ |  |  | ✓ | ✓ | ✓ | ✓ | ✓ | ✓ |  | ✓ | ✓ |
| P747 | 9 |  | ✓ |  | ✓ | ✓ | ✓ |  | ✓ | ✓ | ✓ | ✓ | ✓ |
| P748 | 9 |  | ✓ | ✓ | ✓ | ✓ | ✓ |  | ✓ |  | ✓ | ✓ | ✓ |
| P749 | 9 | ✓ | ✓ | ✓ | ✓ | ✓ | ✓ |  | ✓ |  | ✓ | ✓ |  |
| P750 | 9 |  | ✓ | ✓ |  | ✓ | ✓ | ✓ | ✓ | ✓ | ✓ |  | ✓ |
| P751 | 9 |  | ✓ | ✓ | ✓ | ✓ | ✓ | ✓ | ✓ |  | ✓ |  | ✓ |
| P752 | 9 | ✓ | ✓ | ✓ | ✓ |  | ✓ |  |  | ✓ | ✓ | ✓ | ✓ |
| P753 | 9 | ✓ | ✓ |  |  | ✓ | ✓ |  | ✓ | ✓ | ✓ | ✓ | ✓ |
| P754 | 9 | ✓ | ✓ | ✓ | ✓ |  | ✓ | ✓ |  | ✓ | ✓ |  | ✓ |
| P755 | 9 |  |  | ✓ | ✓ | ✓ |  | ✓ | ✓ | ✓ | ✓ | ✓ | ✓ |
| P756 | 9 |  |  | ✓ | ✓ | ✓ | ✓ | ✓ | ✓ | ✓ |  | ✓ | ✓ |
| P757 | 9 | ✓ |  | ✓ |  | ✓ | ✓ | ✓ |  | ✓ | ✓ | ✓ | ✓ |
| P758 | 9 | ✓ | ✓ |  | ✓ | ✓ | ✓ | ✓ | ✓ |  |  | ✓ | ✓ |
| P759 | 9 | ✓ | ✓ |  | ✓ | ✓ | ✓ | ✓ |  | ✓ | ✓ | ✓ |  |
| P760 | 9 |  | ✓ | ✓ |  | ✓ | ✓ | ✓ | ✓ |  | ✓ | ✓ | ✓ |
| P761 | 9 |  |  | ✓ |  | ✓ | ✓ | ✓ | ✓ | ✓ | ✓ | ✓ | ✓ |
| P762 | 9 |  | ✓ | ✓ | ✓ | ✓ |  | ✓ |  | ✓ | ✓ | ✓ | ✓ |
| P763 | 9 | ✓ | ✓ |  | ✓ | ✓ | ✓ | ✓ |  | ✓ | ✓ |  | ✓ |
| P764 | 9 | ✓ |  | ✓ | ✓ | ✓ |  | ✓ | ✓ | ✓ | ✓ |  | ✓ |
| P765 | 9 | ✓ | ✓ |  | ✓ | ✓ | ✓ |  |  | ✓ | ✓ | ✓ | ✓ |
| P766 | 9 | ✓ | ✓ | ✓ |  | ✓ |  | ✓ | ✓ | ✓ | ✓ | ✓ |  |
| P767 | 9 | ✓ | ✓ | ✓ | ✓ | ✓ |  | ✓ | ✓ |  |  | ✓ | ✓ |
| P768 | 9 | ✓ | ✓ | ✓ | ✓ | ✓ | ✓ | ✓ |  | ✓ |  |  | ✓ |
| P769 | 9 |  |  | ✓ | ✓ | ✓ | ✓ | ✓ |  | ✓ | ✓ | ✓ | ✓ |
| P770 | 9 |  | ✓ |  | ✓ | ✓ | ✓ | ✓ | ✓ | ✓ | ✓ | ✓ |  |
| P771 | 9 | ✓ |  |  | ✓ |  | ✓ | ✓ | ✓ | ✓ | ✓ | ✓ | ✓ |
| P772 | 9 | ✓ | ✓ | ✓ |  |  | ✓ | ✓ | ✓ | ✓ | ✓ | ✓ |  |
| P773 | 9 | ✓ | ✓ | ✓ |  |  | ✓ | ✓ | ✓ |  | ✓ | ✓ | ✓ |
| P774 | 9 |  |  | ✓ | ✓ | ✓ | ✓ | ✓ | ✓ | ✓ | ✓ |  | ✓ |
| P775 | 9 | ✓ | ✓ |  | ✓ | ✓ |  | ✓ | ✓ |  | ✓ | ✓ | ✓ |
| P776 | 9 | ✓ | ✓ |  |  | ✓ | ✓ | ✓ |  | ✓ | ✓ | ✓ | ✓ |
| P777 | 9 | ✓ | ✓ | ✓ |  | ✓ |  | ✓ |  | ✓ | ✓ | ✓ | ✓ |
| P778 | 9 | ✓ |  | ✓ | ✓ |  | ✓ | ✓ | ✓ | ✓ | ✓ |  | ✓ |
| P779 | 9 | ✓ | ✓ | ✓ | ✓ | ✓ |  |  | ✓ | ✓ | ✓ |  | ✓ |
| P780 | 9 | ✓ | ✓ |  | ✓ |  | ✓ | ✓ | ✓ |  | ✓ | ✓ | ✓ |
| P781 | 9 |  | ✓ | ✓ |  | ✓ | ✓ | ✓ |  | ✓ | ✓ | ✓ | ✓ |
| P782 | 9 |  | ✓ | ✓ |  | ✓ | ✓ | ✓ | ✓ | ✓ |  | ✓ | ✓ |
| P783 | 9 |  | ✓ | ✓ | ✓ | ✓ | ✓ | ✓ | ✓ |  | ✓ | ✓ |  |
| P784 | 9 | ✓ | ✓ | ✓ |  | ✓ |  | ✓ | ✓ | ✓ |  | ✓ | ✓ |
| P785 | 9 | ✓ | ✓ | ✓ | ✓ | ✓ |  | ✓ | ✓ | ✓ |  |  | ✓ |
| P786 | 9 | ✓ |  | ✓ |  | ✓ |  | ✓ | ✓ | ✓ | ✓ | ✓ | ✓ |
| P787 | 9 | ✓ | ✓ | ✓ | ✓ |  |  | ✓ | ✓ | ✓ |  | ✓ | ✓ |
| P788 | 9 | ✓ |  | ✓ | ✓ | ✓ | ✓ |  |  | ✓ | ✓ | ✓ | ✓ |
| P789 | 9 |  |  | ✓ | ✓ |  | ✓ | ✓ | ✓ | ✓ | ✓ | ✓ | ✓ |
| P790 | 10 | ✓ | ✓ | ✓ | ✓ |  | ✓ |  | ✓ | ✓ | ✓ | ✓ | ✓ |
| P791 | 10 | ✓ | ✓ |  | ✓ | ✓ | ✓ | ✓ | ✓ | ✓ | ✓ |  | ✓ |
| P792 | 10 | ✓ | ✓ | ✓ | ✓ |  | ✓ | ✓ | ✓ |  | ✓ | ✓ | ✓ |
| P793 | 10 | ✓ | ✓ | ✓ |  | ✓ | ✓ |  | ✓ | ✓ | ✓ | ✓ | ✓ |
| P794 | 10 | ✓ | ✓ | ✓ |  | ✓ | ✓ | ✓ | ✓ | ✓ | ✓ | ✓ |  |
| P795 | 10 | ✓ | ✓ | ✓ | ✓ | ✓ | ✓ | ✓ | ✓ |  | ✓ |  | ✓ |
| P796 | 10 | ✓ | ✓ | ✓ | ✓ | ✓ | ✓ | ✓ |  | ✓ | ✓ |  | ✓ |
| P797 | 10 | ✓ | ✓ | ✓ | ✓ | ✓ | ✓ |  | ✓ | ✓ | ✓ |  | ✓ |
| P798 | 10 | ✓ | ✓ | ✓ | ✓ | ✓ | ✓ |  | ✓ | ✓ |  | ✓ | ✓ |
| P799 | 10 | ✓ | ✓ | ✓ | ✓ | ✓ |  | ✓ | ✓ | ✓ |  | ✓ | ✓ |
| P800 | 10 | ✓ | ✓ | ✓ |  | ✓ | ✓ | ✓ |  | ✓ | ✓ | ✓ | ✓ |
| P801 | 10 |  |  | ✓ | ✓ | ✓ | ✓ | ✓ | ✓ | ✓ | ✓ | ✓ | ✓ |
| P802 | 10 |  | ✓ | ✓ |  | ✓ | ✓ | ✓ | ✓ | ✓ | ✓ | ✓ | ✓ |
| P803 | 10 |  | ✓ |  | ✓ | ✓ | ✓ | ✓ | ✓ | ✓ | ✓ | ✓ | ✓ |
| P804 | 10 | ✓ | ✓ | ✓ | ✓ | ✓ |  |  | ✓ | ✓ | ✓ | ✓ | ✓ |
| P805 | 10 | ✓ | ✓ | ✓ | ✓ | ✓ | ✓ |  |  | ✓ | ✓ | ✓ | ✓ |
| P806 | 10 | ✓ | ✓ |  | ✓ | ✓ | ✓ | ✓ |  | ✓ | ✓ | ✓ | ✓ |
| P807 | 10 | ✓ |  | ✓ | ✓ | ✓ | ✓ | ✓ | ✓ | ✓ | ✓ |  | ✓ |
| P808 | 10 | ✓ | ✓ | ✓ | ✓ |  | ✓ | ✓ |  | ✓ | ✓ | ✓ | ✓ |
| P809 | 10 | ✓ | ✓ | ✓ |  | ✓ | ✓ | ✓ | ✓ | ✓ |  | ✓ | ✓ |
| P810 | 11 | ✓ | ✓ | ✓ | ✓ | ✓ | ✓ | ✓ | ✓ |  | ✓ | ✓ | ✓ |
| P811 | 11 | ✓ | ✓ | ✓ | ✓ | ✓ | ✓ |  | ✓ | ✓ | ✓ | ✓ | ✓ |
| P812 | 11 | ✓ | ✓ | ✓ | ✓ |  | ✓ | ✓ | ✓ | ✓ | ✓ | ✓ | ✓ |
| P813 | 11 | ✓ |  | ✓ | ✓ | ✓ | ✓ | ✓ | ✓ | ✓ | ✓ | ✓ | ✓ |
| P814 | 11 | ✓ | ✓ | ✓ |  | ✓ | ✓ | ✓ | ✓ | ✓ | ✓ | ✓ | ✓ |
| P815 | 11 | ✓ | ✓ | ✓ | ✓ | ✓ |  | ✓ | ✓ | ✓ | ✓ | ✓ | ✓ |
| P816 | 11 | ✓ | ✓ | ✓ | ✓ | ✓ | ✓ | ✓ | ✓ | ✓ | ✓ | ✓ |  |
| P817 | 11 | ✓ | ✓ | ✓ | ✓ | ✓ | ✓ | ✓ | ✓ | ✓ | ✓ |  | ✓ |
| P818 | 11 | ✓ | ✓ |  | ✓ | ✓ | ✓ | ✓ | ✓ | ✓ | ✓ | ✓ | ✓ |
| P819 | 11 | ✓ | ✓ | ✓ | ✓ | ✓ | ✓ | ✓ |  | ✓ | ✓ | ✓ | ✓ |
| P820 | 11 | ✓ | ✓ | ✓ | ✓ | ✓ | ✓ | ✓ | ✓ | ✓ |  | ✓ | ✓ |
| P821 | 11 |  | ✓ | ✓ | ✓ | ✓ | ✓ | ✓ | ✓ | ✓ | ✓ | ✓ | ✓ |

|  | **Full sample**  **(N= 281,729)** | **Training sample**  **(N = 187,819; 66.7%)** | **Validation sample**  **(N= 93,910; 33.3%)** |
| --- | --- | --- | --- |
| **Age** | 57.9 (± 7.8) | 57.9 (± 7.8) | 57.9 (± 7.8) |
| **Sex, n (%)** | 154,085 (54.7%) | 102,392 (54.5%) | 51,693 (55.1%) |
| **Frailty, n (%)** | 16,259 (5.8%) | 10,798 (5.8%) | 5,461 (5.8%) |
| **Clinical parameters** |  |  |  |
| BMI | 27.9 (± 5.0) | 27.9 (± 5.0) | 28.0 (± 5.0) |
| Total cholesterol | 5.6 (± 1.2) | 5.6 (± 1.2) | 5.6 (± 1.2) |
| LDL cholesterol | 3.5 (± 0.9) | 3.5 (± 0.9) | 3.5 (± 0.9) |
| HDL cholesterol | 1.4 (± 0.4) | 1.4 (± 0.4) | 1.4 (± 0.4) |
| Triglycerides | 1.8 (± 1.0) | 1.8 (± 1.0) | 1.8 (± 1.0) |
| ALAT | 24.0 (± 14.6) | 24.1 (± 14.6) | 24.0 (± 14.5) |
| ASAT | 26.6 (± 11.0) | 26.6 (± 11.0) | 26.6 (± 10.9) |
| GGT | 39.7 (± 46.2) | 39.8 (± 46.0) | 39.7 (± 46.4) |
| Creatinine | 0.8 (± 0.2) | 0.8 (± 0.2) | 0.8 (± 0.3) |
| eGFR | 89.3 (± 13.8) | 89.3 (± 13.8) | 89.3 (± 13.9) |
| HbA1c | 36.8 (± 7.4) | 36.8 (± 7.4) | 36.8 (± 7.5) |
| Glucose | 5.2 (± 1.4) | 5.2 (± 1.4) | 5.2 (± 1.4) |
| **Health profile, n (%)** |  |  |  |
| 3-year mortality | 3,047 (1.1%) | 2,031 (1.1%) | 1,016 (1.1%) |
| 10-year mortality | 17,164 (6.1%) | 11,367 (6.1%) | 5,797 (6.2%) |
| Polypharmacy | 73,421 (26.1%) | 48,950 (26.1%) | 24,471 (26.1%) |
| Fall during the past year | 61,667 (21.9%) | 41,151 (21.9%) | 20,516 (21.8%) |
| Poor rated overall health | 17,696 (6.4%) | 11,934 (6.4%) | 5,762 (6.1%) |

**Supplementary Table 3. Participants characteristics.**

Statistics are mean (± SD) unless specified otherwise.

Abbreviations: BMI: Body mass index; SD: standard deviation; LDL: low density lipoprotein; HDL: high density lipoprotein; ALAT: Alanine aminotransferase; ASAT: aspartate aminotransferase; GGT: gamma-glutamyl transferase; eGFR: estimated glomerular filtration rate; HbA1c : glycated hemoglobin A1C.

**Supplementary Table 4. Gain, lift and number of cases stratified by top k%, assessed in validation sample (N= 93,910).**

| **Top k%^A^** | **Participants** | **Frail participants** | **% frail in top k%^B^** | **Gain^C^** | **Lift** |
| --- | --- | --- | --- | --- | --- |
| **10%** | 9,391 | 2,080 | 22.1% | 38.1% | 3.81 |
| **20%** | 18,782 | 3,039 | 16.2% | 55.6% | 2.78 |
| **30%** | 28,173 | 3,665 | 13.0% | 67.1% | 2.24 |
| **50%** | 46,955 | 4,535 | 9.7% | 83.0% | 1.66 |
| **75%** | 70,433 | 5,149 | 7.3% | 94.3% | 1.26 |
| **Complete sample (100%)** | 93,910 | 5,461 | 5.8% | 100% | 1 |

^A^ Top k% refers to the percentage of participants, sorted by decreasing predicted frailty, included in the stratum.
^B^ denominator is number of participants in the top k% (column 2)
^C^ denominator is total number of participants with frailty in complete sample (n = 5,461)

**Supplementary Table 5. Summary statistics of distribution of normalized gain, gain and lift at top k% across 3,284 alternative assessments.**

|  | **Normalized gain** | **Top 10%** | | **Top 20%** | | **Top 30%** | |
| --- | --- | --- | --- | --- | --- | --- | --- |
|  |  | **Gain** | **Lift** | **Gain** | **Lift** | **Gain** | **Lift** |
| Median | 39.4% | 29.6% | 3.0 | 46.0% | 2.3 | 57.9% | 1.9 |
| Q1 | 34.3% | 25.5% | 2.6 | 41.0% | 2.1 | 53.2% | 1.8 |
| Q3 | 42.9% | 32.2% | 3.2 | 48.9% | 2.5 | 60.6% | 2.0 |
| IQR | 8.6% | 6.2% | 0.6 | 7.9% | 0.4 | 7.4% | 0.2 |
| P5 | 28.1% | 22.6% | 2.3 | 37.0% | 1.9 | 48.8% | 1.6 |
| P95 | 45.6% | 33.9% | 3.4 | 50.7% | 2.5 | 62.4% | 2.1 |

Abbreviation: Q1: 1^st^ quartile; Q3: 3^rd^ quartile; IQR: interquartile range; P5: 5^th^ percentile; P95: 95^th^ percentile.

**Supplementary Table 6.** **Determinants of variability in gain at top 10 and top 30%.**

| **Predictor** | **Unique variance explained (Part R²)** | | **Total variance associated with predictor (Inclusive R²)** | |
| --- | --- | --- | --- | --- |
|  | **Top 10%** | **Top 30%** | **Top 10%** | **Top 30%** |
| Number of clinical measures | 6.7% (1.4; 12.0) | 7.8% (3.0; 13.5) | 18.5% (13.7; 22.9) | 22.1% (17.4; 26.8) |
| Panel cost | 0.2% (0.0; 0.6) | 0.2% (0.0; 6.2) | 12.1% (8.6; 16.1) | 14.5% (10.2; 18.6) |
| Age group | 0.0% (0.0; 0.6) | 0.2% (0.0; 6.2) | 0.0% (0.0; 0.1) | 0.2% (0.1; 0.2) |
| Disease list choice | 0.2% (0.0; 0.6) | 0.0% (0.0; 6.0) | 0.2% (0.1; 0.3) | 0.0% (0.0; 0.0) |
| Marginal R² | **Top 10%:** 91.2%; **Top 30%:** 93.7% | | | |
| Conditional R² | **Top 10%:** 18.9%; **Top 30%:** 22.5% | | | |
| Random intercept variance attributable  to clinical measure | **Top 10%:** 72.3%; **Top 30%:** 71.2% | | | |

**Supplementary Table 7. Difference in median gain in combination with or without a given clinical parameter.**

| **Panel size** | **Clinical parameter** | **Difference in median gain** | **Median gain with** | **Median gain without** | **Number of panels with** | **Number of panels without** |
| --- | --- | --- | --- | --- | --- | --- |
| 3 | BMI | 6.3 | 46.1 | 39.8 | 32 | 144 |
| 3 | Total cholesterol | 0.9 | 40.9 | 40.0 | 40 | 136 |
| 3 | LDL cholesterol | -0.7 | 39.9 | 40.5 | 24 | 152 |
| 3 | HDL cholesterol | -0.7 | 40.0 | 40.6 | 40 | 136 |
| 3 | Triglycerides | -0.7 | 40.0 | 40.7 | 44 | 132 |
| 3 | ALAT | -3.4 | 38.5 | 41.9 | 44 | 132 |
| 3 | ASAT | 6.8 | 46.6 | 39.8 | 32 | 144 |
| 3 | GGT | 1.8 | 41.8 | 40.0 | 44 | 132 |
| 3 | Creatinine | -1.0 | 39.8 | 40.7 | 64 | 112 |
| 3 | eGFR | 0.8 | 40.7 | 40.0 | 48 | 128 |
| 3 | HbA1c | -1.3 | 39.4 | 40.7 | 60 | 116 |
| 3 | Glucose | -1.8 | 38.9 | 40.6 | 56 | 120 |
| 4 | BMI | 0.2 | 41.6 | 41.4 | 156 | 240 |
| 4 | Total cholesterol | 0.8 | 42.0 | 41.3 | 120 | 276 |
| 4 | LDL cholesterol | -1.0 | 40.9 | 41.8 | 128 | 268 |
| 4 | HDL cholesterol | -0.5 | 41.1 | 41.6 | 152 | 244 |
| 4 | Triglycerides | 1.6 | 42.7 | 41.1 | 108 | 288 |
| 4 | ALAT | -2.6 | 40.2 | 42.8 | 144 | 252 |
| 4 | ASAT | 1.3 | 42.4 | 41.1 | 116 | 280 |
| 4 | GGT | -0.5 | 41.1 | 41.6 | 160 | 236 |
| 4 | Creatinine | 0.0 | 41.4 | 41.4 | 132 | 264 |
| 4 | eGFR | 0.1 | 41.4 | 41.3 | 148 | 248 |
| 4 | HbA1c | 1.6 | 42.7 | 41.1 | 112 | 284 |
| 4 | Glucose | 4.5 | 45.6 | 41.1 | 108 | 288 |
| 5 | BMI | -0.4 | 40.8 | 41.2 | 232 | 404 |
| 5 | Total cholesterol | -1.2 | 40.4 | 41.6 | 264 | 372 |
| 5 | LDL cholesterol | 0.4 | 41.2 | 40.9 | 260 | 376 |
| 5 | HDL cholesterol | -1.2 | 40.4 | 41.6 | 292 | 344 |
| 5 | Triglycerides | 0.4 | 41.2 | 40.8 | 244 | 392 |
| 5 | ALAT | -0.2 | 40.9 | 41.1 | 252 | 384 |
| 5 | ASAT | 0.3 | 41.2 | 40.9 | 248 | 388 |
| 5 | GGT | 0.8 | 41.4 | 40.6 | 280 | 356 |
| 5 | Creatinine | 0.0 | 41.0 | 41.0 | 284 | 352 |
| 5 | eGFR | 1.2 | 41.9 | 40.7 | 244 | 392 |
| 5 | HbA1c | 0.1 | 41.1 | 41.0 | 284 | 352 |
| 5 | Glucose | 0.2 | 41.2 | 41.0 | 296 | 340 |
| 6 | BMI | -0.3 | 46.6 | 46.9 | 380 | 360 |
| 6 | Total cholesterol | -0.3 | 46.6 | 46.9 | 388 | 352 |
| 6 | LDL cholesterol | -1.8 | 45.2 | 46.9 | 368 | 372 |
| 6 | HDL cholesterol | 1.8 | 47.4 | 45.6 | 344 | 396 |
| 6 | Triglycerides | -1.4 | 45.9 | 47.2 | 396 | 344 |
| 6 | ALAT | 3.2 | 47.5 | 44.3 | 396 | 344 |
| 6 | ASAT | -0.3 | 46.6 | 46.9 | 404 | 336 |
| 6 | GGT | 1.0 | 47.0 | 46.0 | 360 | 380 |
| 6 | Creatinine | -2.9 | 44.4 | 47.3 | 324 | 416 |
| 6 | eGFR | 3.1 | 47.5 | 44.4 | 324 | 416 |
| 6 | HbA1c | 0.1 | 46.8 | 46.7 | 392 | 348 |
| 6 | Glucose | -3.1 | 44.4 | 47.5 | 364 | 376 |
| 7 | BMI | 0.6 | 48.3 | 47.7 | 384 | 252 |
| 7 | Total cholesterol | 0.3 | 48.2 | 47.8 | 372 | 264 |
| 7 | LDL cholesterol | 0.4 | 48.2 | 47.8 | 340 | 296 |
| 7 | HDL cholesterol | 0.1 | 48.0 | 47.9 | 364 | 272 |
| 7 | Triglycerides | 0.1 | 48.0 | 47.9 | 368 | 268 |
| 7 | ALAT | 2.1 | 48.3 | 46.2 | 392 | 244 |
| 7 | ASAT | -1.3 | 47.4 | 48.7 | 400 | 236 |
| 7 | GGT | 0.2 | 48.0 | 47.8 | 404 | 232 |
| 7 | Creatinine | -0.5 | 47.7 | 48.3 | 344 | 292 |
| 7 | eGFR | 0.7 | 48.2 | 47.5 | 384 | 252 |
| 7 | HbA1c | -1.3 | 47.1 | 48.4 | 348 | 288 |
| 7 | Glucose | -0.1 | 48.0 | 48.1 | 352 | 284 |
| 8 | BMI | -0.6 | 47.6 | 48.2 | 252 | 144 |
| 8 | Total cholesterol | -0.7 | 47.5 | 48.2 | 256 | 140 |
| 8 | LDL cholesterol | 0.3 | 47.9 | 47.6 | 284 | 112 |
| 8 | HDL cholesterol | -0.2 | 47.7 | 47.9 | 264 | 132 |
| 8 | Triglycerides | 0.9 | 48.2 | 47.3 | 244 | 152 |
| 8 | ALAT | 0.4 | 47.9 | 47.5 | 268 | 128 |
| 8 | ASAT | -0.5 | 47.6 | 48.1 | 244 | 152 |
| 8 | GGT | 0.2 | 47.9 | 47.7 | 288 | 108 |
| 8 | Creatinine | -0.7 | 47.5 | 48.2 | 252 | 144 |
| 8 | eGFR | 0.5 | 47.9 | 47.4 | 260 | 136 |
| 8 | HbA1c | 0.8 | 48.0 | 47.3 | 268 | 128 |
| 8 | Glucose | 1.0 | 48.0 | 47.0 | 288 | 108 |
| 9 | BMI | 6.1 | 50.0 | 44.0 | 112 | 64 |
| 9 | Total cholesterol | 0.1 | 48.5 | 48.5 | 124 | 52 |
| 9 | LDL cholesterol | -1.2 | 48.2 | 49.4 | 128 | 48 |
| 9 | HDL cholesterol | 0.1 | 48.6 | 48.5 | 120 | 56 |
| 9 | Triglycerides | -2.0 | 48.2 | 50.2 | 140 | 36 |
| 9 | ALAT | -1.8 | 48.1 | 49.9 | 128 | 48 |
| 9 | ASAT | -0.2 | 48.5 | 48.7 | 144 | 32 |
| 9 | GGT | 0.3 | 48.7 | 48.4 | 124 | 52 |
| 9 | Creatinine | -0.5 | 48.5 | 48.9 | 136 | 40 |
| 9 | eGFR | 0.2 | 48.5 | 48.4 | 136 | 40 |
| 9 | HbA1c | -0.2 | 48.5 | 48.6 | 152 | 24 |
| 9 | Glucose | -1.3 | 48.3 | 49.6 | 140 | 36 |
| 10 | BMI | 5.8 | 50.2 | 44.4 | 68 | 12 |
| 10 | Total cholesterol | 2.3 | 50.0 | 47.7 | 72 | 8 |
| 10 | LDL cholesterol | 1.1 | 50.0 | 48.9 | 68 | 12 |
| 10 | HDL cholesterol | -0.3 | 49.8 | 50.0 | 60 | 20 |
| 10 | Triglycerides | -0.8 | 49.7 | 50.5 | 68 | 12 |
| 10 | ALAT | -0.5 | 49.8 | 50.2 | 72 | 8 |
| 10 | ASAT | -0.5 | 49.7 | 50.1 | 56 | 24 |
| 10 | GGT | 0.9 | 50.0 | 49.2 | 60 | 20 |
| 10 | Creatinine | -1.1 | 49.7 | 50.8 | 72 | 8 |
| 10 | eGFR | 0.3 | 50.0 | 49.7 | 60 | 20 |
| 10 | HbA1c | -0.7 | 49.8 | 50.4 | 76 | 4 |
| 10 | Glucose | -0.9 | 49.7 | 50.5 | 68 | 12 |
| 11 | BMI | 6.1 | 50.4 | 44.4 | 44 | 4 |
| 11 | Total cholesterol | -0.8 | 50.2 | 51.0 | 44 | 4 |
| 11 | LDL cholesterol | -0.3 | 50.4 | 50.6 | 44 | 4 |
| 11 | HDL cholesterol | -0.1 | 50.4 | 50.5 | 44 | 4 |
| 11 | Triglycerides | -0.2 | 50.4 | 50.6 | 44 | 4 |
| 11 | ALAT | -0.2 | 50.4 | 50.5 | 44 | 4 |
| 11 | ASAT | -0.4 | 50.4 | 50.8 | 44 | 4 |
| 11 | GGT | 1.3 | 50.4 | 49.1 | 44 | 4 |
| 11 | Creatinine | -0.7 | 50.2 | 50.9 | 44 | 4 |
| 11 | eGFR | 0.5 | 50.4 | 49.9 | 44 | 4 |
| 11 | HbA1c | -0.3 | 50.2 | 50.5 | 44 | 4 |
| 11 | Glucose | -0.8 | 50.2 | 51.0 | 44 | 4 |

Abbreviations: BMI: Body mass index; LDL: low density lipoprotein; HDL: high density lipoprotein; ALAT: Alanine aminotransferase; ASAT: aspartate aminotransferase; GGT: gamma-glutamyl transferase; eGFR: estimated glomerular filtration rate; HbA1c : glycated hemoglobin A1C.

**Supplementary Table 8. Gain at top 20% and normalized gain in full sample, by age groups (defined by quartile) or by patient characteristics (presence/absence metabolic or cardiometabolic diseases).**

|  | **Gain at top 20%** | **Normalized gain** |
| --- | --- | --- |
| **Full sample** | 55.6% | 53.4% |
| **Age groups** | | |
| [38–52) | 56.3% | 51.4% |
| [52–60) | 55.2% | 54.0% |
| [60–64) | 54.1% | 52.6% |
| [64–71] | 56.4% | 54.2% |
| **Any metabolic disease** | | |
| Any | 53.0% | 54.7% |
| None | 53.2% | 50.4% |
| **Cardiometabolic disease** | | |
| Any | 53.2% | 51.9% |
| None | 51.5% | 47.8% |

**Supplementary Figure 1: Flow chart of UK Biobank participants.**

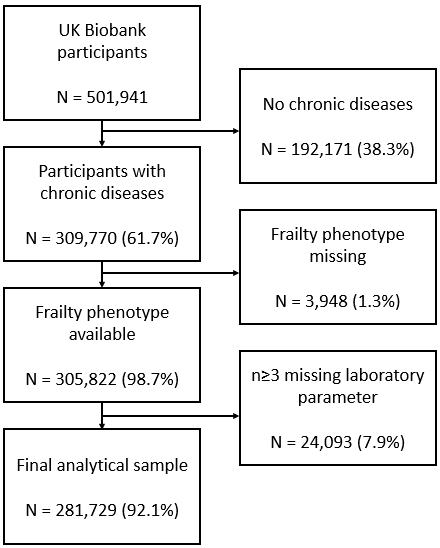

**Supplementary Figure 2. Summary statistics of distribution of gain at top 20% by length of combination and cost of combination across 3,284 alternative model settings.**

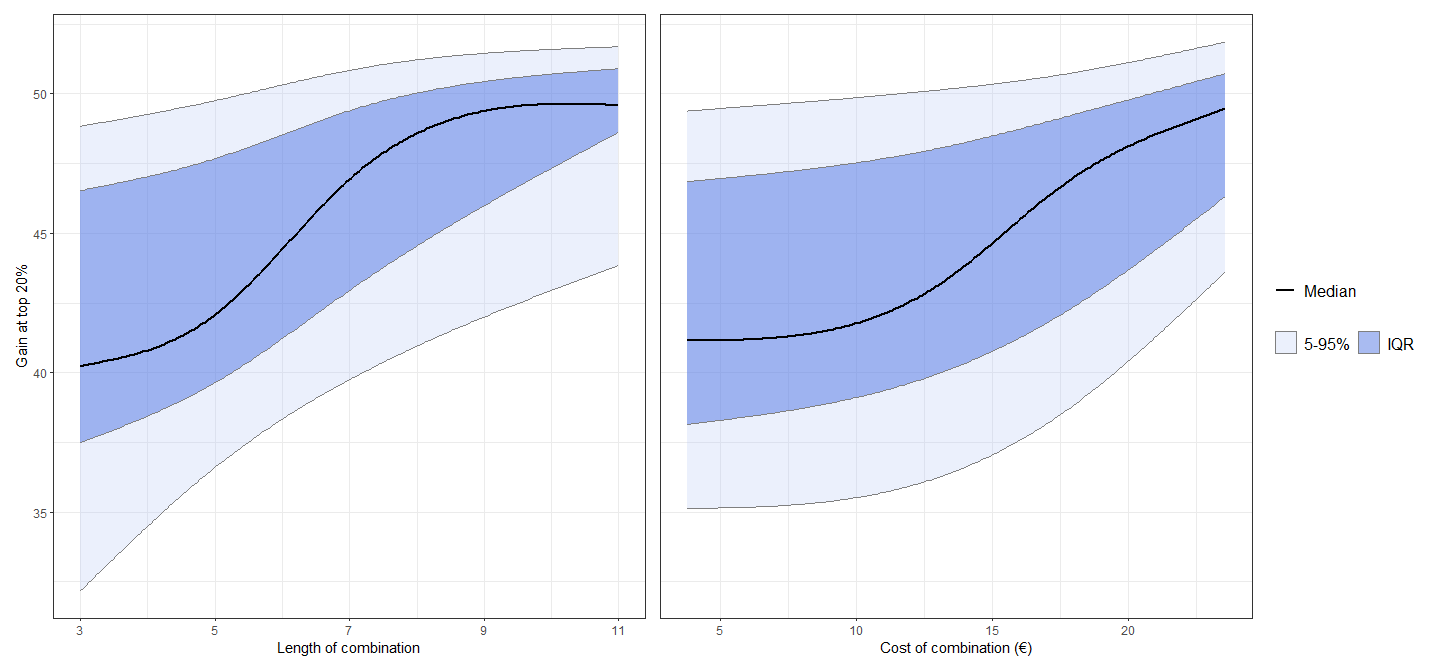

**Supplementary Figure 3. Gain curve, gain at top 20% and normalized gain in age subgroups defined by quartiles.**

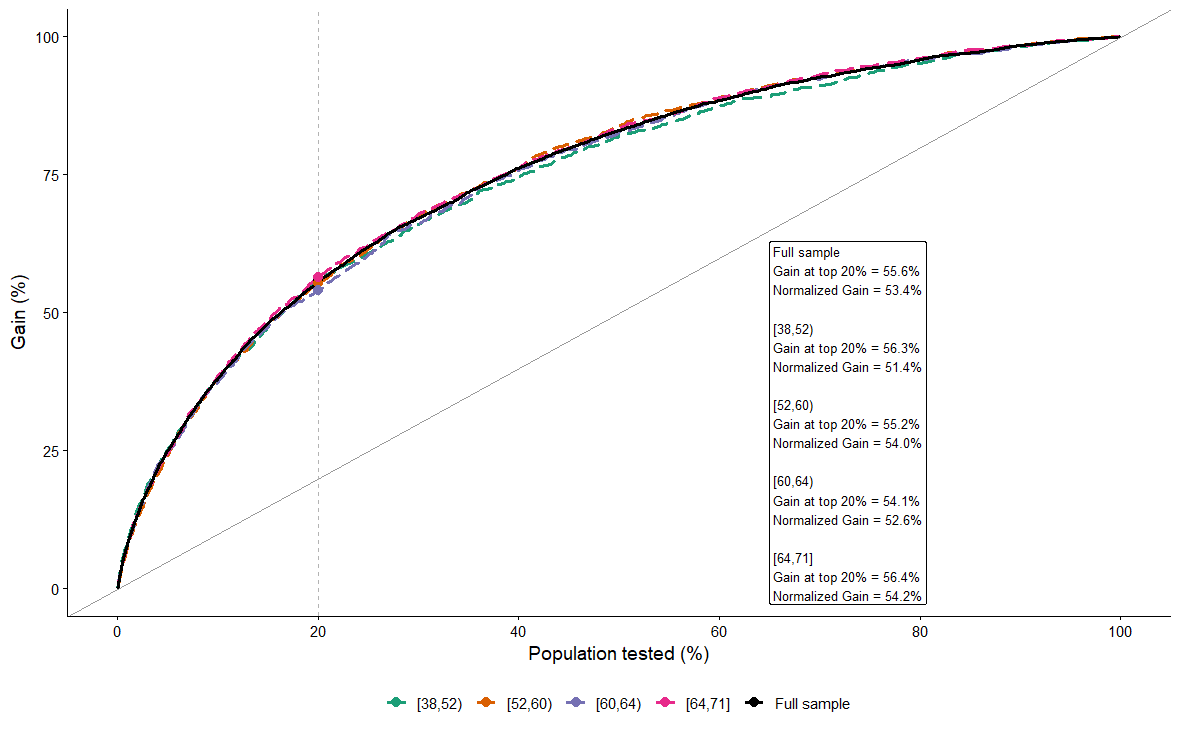

**Supplementary Figure 4. Gain curve, gain at top 20% and normalized gain in patient subgroups defined by presence/absence of metabolic or cardiometabolic diseases.**

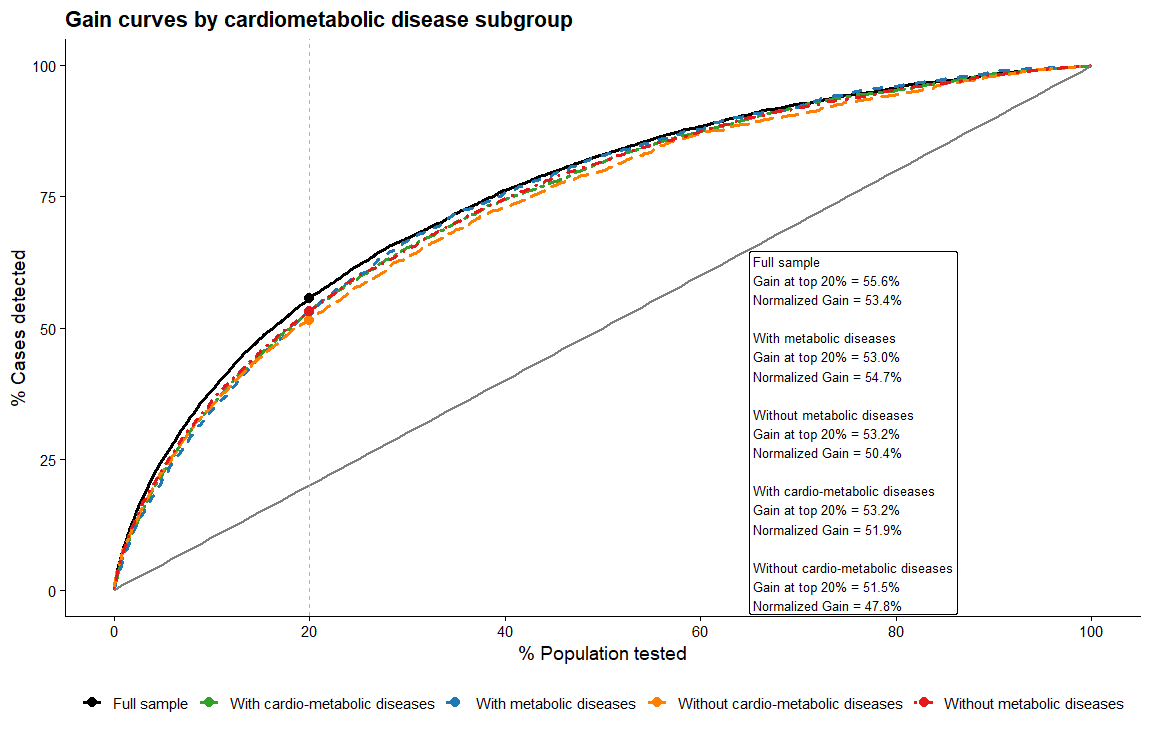

**Supplementary method 1.**

A. **Measurement of routine clinical measures.**

The following measurement procedures were applied:

- BMI: defined using measured weight and height.
- Total cholesterol, triglycerides, HDL: enzymatically by use of a Beckmann Coulter AU5800
- ALAT, ASAT, GGT: Enzymatic Rate
- LDL cholesterol: Enzymatic Selective Protection by use of a Beckmann Coulter AU5800
- HbA1c: high performance liquid chromatography on a Bio-Rad VARIANT II Turbo
- Glucose: hexokinase analysis on a Beckman Coulter AU5800.
- eGFR: calculated using the 2009 CKD-EPI creatinine equation (without the race coefficient).

**Abbreviations:** BMI: Body mass index; SD: standard deviation; LDL: low density lipoprotein; HDL: high density lipoprotein; ALAT: Alanine aminotransferase; ASAT: aspartate aminotransferase; GGT: gamma-glutamyl transferase; eGFR: estimated glomerular filtration rate; HbA1c : glycated hemoglobin A1C.

**Reference:** UK Biobank. UK Biobank Showcase. [https://biobank.ndph.ox.ac.uk/showcase/search.cgi. Accessed 12 Nov 2025](https://biobank.ndph.ox.ac.uk/showcase/search.cgi.%20Accessed%2012%20Nov%202025).

B. **Cost calculation**

Cost calculation was estimated using publicly available reimbursement data from the French National Health Insurance system. We used the information from the 2025 database.

For a given routine clinical measure, cost was calculated as the total amount reimbursed during 2025, divided by the number of procedures performed.

Body mass index has a cost of 0, given the availability of its measuring instruments at home.

For measures derived from other parameters (eGFR, HDL cholesterol): the cost is 0 if, in the combination under consideration, the input parameters are already present; otherwise, the cost is equal to the cost of the parameters required to calculate it.

| **Routine clinical measure** | **Estimated cost (€)** | **Combined estimated cost (€)** |
| --- | --- | --- |
| Total cholesterol | 1.3 | 1.3 |
| HDL cholesterol | 5.8 | 5.8 |
| LDL cholesterol | 4.5 | 4.5 |
| Triglycerides | 1.5 | 1.5 |
| eGFR | 7.5 | 7.5 |
| Creatinine | 7.5 | 0 |
| ASAT | 1.5 | 1.5 |
| ALAT | 1.5 | 0 |
| GGT | 1.3 | 1.3 |
| HbA1c | 3.3 | 3.3 |
| Glucose | 1.3 | 1.3 |
| BMI | 0 | 0 |
| Total | 37 | 28 |

**Abbreviations:** BMI: Body mass index; SD: standard deviation; LDL: low density lipoprotein; HDL: high density lipoprotein; ALAT: Alanine aminotransferase; ASAT: aspartate aminotransferase; GGT: gamma-glutamyl transferase; eGFR: estimated glomerular filtration rate; HbA1c : glycated hemoglobin A1C.

**Reference:** [Actes de biologie médicale par type de prescripteur - Biol'AM - 2012 à 2026 | L'Assurance Maladie](https://www.assurance-maladie.ameli.fr/etudes-et-donnees/actes-biologie-medicale-type-prescripteur-biolam)

**Supplementary method 2: prediction modelling strategy.**

0. **Summary of modelling strategy**

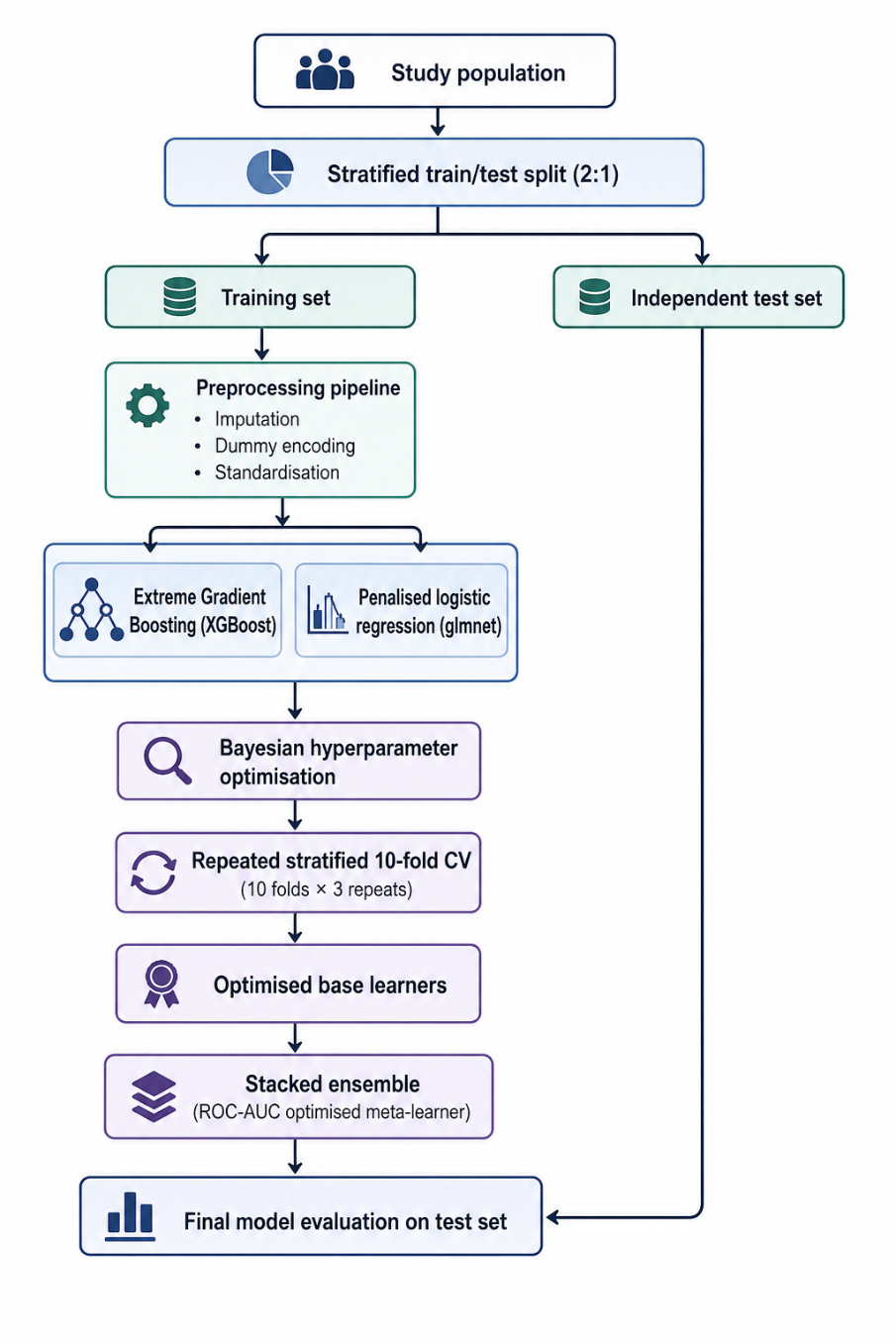

**Overview of the modelling strategy.** The dataset was split into training and validation sets using stratified sampling. Within the training set, preprocessing, hyperparameter optimisation and model selection were performed using repeated stratified 10-fold cross-validation. Extreme gradient boosting and penalised logistic regression were combined using a stacked ensemble optimised for ROC-AUC. Final predictive performance was assessed on the held-out validation set.

1. **Data preparation**

Candidate predictors included demographic characteristics and routine biochemical markers (sex, age, Body Mass Index (BMI), total cholesterol, low-density lipoprotein (LDL) cholesterol, high-density lipoprotein (HDL) cholesterol, triglycerides, alanine aminotransferase (ALAT), aspartate aminotransferase (ASAT), gamma-glutamyl transferase (GGT), creatinine, estimated glomerular filtration rate (eGFR), glycated haemoglobin A1c (HbA1c), glucose). The outcome was a binary frailty classification. Continuous predictors with missing values were imputed using median imputation, categorical variables were dummy encoded, and continuous predictors were standardized. Preprocessing parameters (including imputation values and scaling statistics) were estimated using only the analysis subset within each cross-validation resample and subsequently applied to the corresponding assessment subset.

2. **Training and valiation datasets**

The dataset was randomly divided into a training set (two-thirds of participants) and an independent validation set (one-third) using stratified sampling to preserve the proportion of frailty cases.

3. **Internal resampling**

Model development was performed exclusively within the training set using repeated stratified 10-fold cross-validation (10 folds, three repeats). This resampling strategy was used for hyperparameter optimization and internal performance estimation while accounting for outcome imbalance.

4. **Base learning algorithms**

Two complementary prediction algorithms were considered. Extreme gradient boosting (XGBoost) was used to capture nonlinear relationships and higher-order interactions, whereas penalised logistic regression provided a sparse linear classifier with regularization. Both models were developed within the same preprocessing pipeline.

| **Model** | **Hyperparameter** | **Search range** |
| --- | --- | --- |
| XGBoost | Number of trees | 200–600 |
|  | Maximum tree depth | 3–6 |
|  | Learning rate | 0.01–0.10 |
|  | Minimum observations per terminal node | 5–20 |
| Penalised logistic regression | Penalty (λ) | 10⁻⁴–1 (log scale) |
|  | Elastic-net mixing parameter (α) | 0–1 |

5. **Hyperparameter optimization**

Hyperparameters were optimized separately for each base learner using Bayesian optimization. An initial space-filling design of 10 hyperparameter combinations was evaluated, followed by 20 Bayesian optimization iterations. Each candidate configuration was evaluated using repeated stratified 10-fold cross-validation (10 folds, three repeats), yielding performance estimates across 30 resamples.

6. **Stacked ensemble**

Predictions from the tuned base learners were combined using stacked generalization. Cross-validated out-of-fold predictions from each candidate model were used to train a penalized meta-learner, which estimated the optimal combination of base learners by maximizing ROC-AUC. The final ensemble was then refitted using the complete training dataset.

7. **Independent evaluation and comparison with individual models**

Final model performance was assessed on the independent validation set.

**Supplementary Methods 3. Gain and Lift Metrics for Screening Prioritization**

0. **Overall rational**

The intended use of the prediction model was to prioritize individuals for frailty screening rather than to establish a diagnosis. Consequently, model performance was evaluated using gain and lift metrics, which quantify how effectively the model concentrates individuals with frailty within the highest-risk segments of the population.

Gain and lift metrics directly address a practical screening question: ***If only a proportion of the population can be screened, what proportion of frail individuals would be identified?***

1. **Gain**

Participants were ranked according to their predicted probability of frailty, from highest to lowest risk. For a given screening thresholds (e.g., the highest-risk 10%, 20%, or 30% of participants), gain was defined as the cumulative proportion of all frail individuals captured within that subgroup.

For example, if 40% of all frail individuals were contained within the 20% of participants with the highest predicted risk, the gain at 20% screening coverage would be 40%.

2. **Area under the gain curve**

A gain curve was constructed by plotting the cumulative proportion of frail individuals identified against the cumulative proportion of the population screened. Three reference curves can be considered:

- ***Random selection***, in which the proportion of frail individuals identified increases proportionally with the proportion screened;
- ***Model-based selection***, corresponding to the prediction model under evaluation;
- ***Optimal selection***, representing a theoretical upper bound in which all frail individuals are screened before any non-frail individuals.

A model with greater screening utility produces a gain curve that lies further above the random-selection curve and closer to the optimal selection curve.

To provide an overall assessment across all screening thresholds, the area under the gain curve was calculated and normalized relative to the areas obtained under random and optimal selection. This normalized gain statistic ranges from 0 to 100%, where:

- 0 corresponds to performance equivalent to random selection;
- 100% corresponds to the optimal selection of frail individuals.

3. **Lift**

Lift quantifies the improvement achieved by model-based screening compared with random selection. It is calculated as the ratio between the observed gain and the gain expected under random sampling at the same screening thresholds.

For example, if screening the highest-risk 20% of participants identifies 40% of frail individuals, whereas random selection would identify 20%, the lift at 20% is 2.0. This indicates that the model identifies frail individuals twice as efficiently as random screening.

A lift value of:

- 1 indicates no improvement over random selection;
- > 1 indicates improved targeting efficiency;

The larger the lift value, the greater the concentration of frail individuals within the selected subgroup.

4. **Reported Screening Thresholds**

Gain and lift values were reported for the top 10%, 20%, 30%, 50%, 75%, and 100% of participants ranked according to predicted frailty probability. These thresholds were chosen to reflect a range of potential screening capacities, from highly targeted programs to population-wide screening.

**Supplementary Methods 4. Adaptation of main pipeline for subgroup analyses.**

To ensure reasonable computation time across the 3,284 subgroups, we applied the following modifications of the parameters in the resampling and hyperparameters search strategies while maintaining the same steps.

|  | **Main analysis** | **Subgroup analysis** |
| --- | --- | --- |
| **Resampling** | Repeated (3) CV (10) | CV (3) |
| **Hyperparameter grid definition** | Bayesian search grid (10) | Grid space filling (5) |

**Supplementary methods 5. Count of possible combinations, those selected, and those evaluated.**

The table below lists the possible combinations and those included in the analyses. The qualitative combinations are listed in **Supplementary Table 2**.

| **Panel size** | **Total combinations** | **Retained combinations†** | **Stacked models evaluated‡** |
| --- | --- | --- | --- |
| 3 | 220 | 44 | 176 |
| 4 | 495 | 99 | 396 |
| 5 | 792 | 159 | 636 |
| 6 | 924 | 185 | 740 |
| 7 | 792 | 159 | 636 |
| 8 | 495 | 99 | 396 |
| 9 | 220 | 44 | 176 |
| 10 | 66 | 20 | 80 |
| 11 | 12 | 12 | 48 |
| **Total from 3 to 11** | 4,016 | 821 | 3,284 |

† Twenty percent of all possible combinations were randomly sampled. A minimum of 20 combinations was retained for each panel size. When fewer than 20 possible combinations existed, all combinations were included.

‡ Four stacked models were evaluated for each retained clinical panel combination (corresponding to the combinations of age stratum and chronic disease definition).
